# Cost-effectiveness of use of 20-valent pneumococcal conjugate vaccine among adults in Germany

**DOI:** 10.1101/2023.02.16.23286052

**Authors:** Felicitas Kühne, Katharina Achtert, Franziska Püschner, Dominika Urbanski-Rini, Juliane Schiller, Ernestine Mahar, Josephine Friedrich, Mark Atwood, Ralf Sprenger, Jeffrey Vietri, Christof von Eiff, Christian Theilacker

## Abstract

**Objectives:** Despite national recommendations for use of pneumococcal vaccines, rates of community-acquired pneumonia (CAP) and invasive pneumococcal disease (IPD) remain high in Germany. New pneumococcal conjugate vaccines (PCVs) with expanded coverage have the potential to reduce the pneumococcal disease burden among adults.

**Methods:** Using a Markov model, we evaluated the lifetime outcomes/costs comparing 20-valent PCV (PCV20) with standard of care (SC) vaccination for prevention of CAP and IPD among adults aged ≥60 years and at-risk adults aged 18-59 years in Germany. PCV20 also was compared with sequential vaccination with 15-valent PCV (PCV15) followed by (→) PPSV23 in a scenario analysis.

**Results:** Over the course of a lifetime (82 years), use of PCV20 vs. SC would prevent 54,333 hospitalizations, 26,368 outpatient CAP cases, 10,946 disease-related deaths yield 74,694 additional life-years (LYs), while lowering total medical costs by 363.2M €. PCV20 remained cost saving (i.e., dominant) versus SC even in numerous sensitivity analyses. In the scenario analysis, PCV20 also showed to be cost-saving compared to using PCV15→PPSV23.

**Conclusions:** One dose of PCV20 among adults aged ≥60 years and adults aged 18-59 years with moderate- and high-risk conditions would substantially reduce pneumococcal disease, save lives, and be cost saving compared with SC.

**HIGHLIGHTS:**

- Pneumococcal disease causes significant morbidity and mortality among adults in Germany
- New, higher valent vaccines have the potential to reduce disease burden and associated costs in vulnerable populations
- ver a lifetime, 20-valent pneumococcal conjugate vaccine was found to be cost-saving compared with current standard of care for pneumococcal disease prevention among adults in Germany

## 1 INTRODUCTION

*Streptococcus pneumoniae* is a major cause of invasive (IPD) and non-invasive diseases and associated with high rates of hospitalizations and premature death, especially among older adults [1]. Community-acquired pneumonia (CAP) is the most common manifestation with more than 250,000 hospitalizations and 200,000 outpatient visits annually among adults in Germany [1–3]. Interpretation of IPD trends in adults recorded by Germany’s national reference center are difficult due to improvements in disease reporting over time [4]. However, several European countries with robust IPD surveillance systems have reported an increase in IPD incidence among older adults in the 4 years prior to the COVID-19 pandemic [5–7].

Three vaccines have been available in Germany until recently to prevent pneumococcal disease: a 23-valent pneumococcal polysaccharide vaccine (PPSV23), a 13-valent pneumococcal conjugate vaccine (PCV13), and 10-valent PCV (PCV10) [8]. The national Standing Committee on Vaccination (STIKO) recommends PCVs for children aged ≤24 months (2+1 schedule for term infants), and sequentially administered PCV13 and PPSV23 (PCV13→PPSV23) for children and adolescents aged 2-15 years with underlying medical conditions [8]. A single dose of PPSV23 is recommended for immunocompetent individuals aged ≥16 years with chronic medical conditions and for all immunocompetent adults aged ≥60 years. A single dose of PCV13 followed by PPSV23 (with at least 6-month interval) is recommended for all adults aged ≥16 years with immunocompromising conditions [8]. According to STIKO recommendations, revaccination with PPSV23 after at least 6 years may be considered for individuals aged ≥16 years who have previously received PPSV23 [8]. Currently STIKO is assessing the risk benefit of higher-valent PCVs and respective recommendation is not published yet.

As a result of the indirect effects of paediatric PCV vaccination, the burden of IPD due to serotypes included in 10- and 13-valent PCVs has decreased among adults reaching a steady state in the last years [7]. Nevertheless, the overall burden of pneumococcal disease among adults has increased, highlighting the need for vaccines with expanded serotype coverage [7]. Next generation PCVs have the potential to further reduce the burden of pneumococcal disease in Germany [9].

Meanwhile, two higher-valent PCVs - a 20-valent PCV (PCV20) and a 15-valent PCV (PCV15) - that include the serotypes contained in PCV13 as well as additional epidemiologically important serotypes have been approved for use in adults by the European Medicines Agency (EMA)[10–12]. PCV20 contains the serotypes in PCV15 (1, 3, 4, 5, 6A, 6B, 7F, 9V, 14, 18C, 19A, 19F, 23F, 22F, 33F) plus five additional serotypes (8, 10A, 11A, 12F, 15B) [13,14]. Although nineteen of the twenty serotypes in PCV20 are also present in PPSV23, PCV20 is expected to provide more robust protection due to its T-cell dependent immune response, generation of immunologic memory and induction of an antigenically broader and more durable antibody response [15,16]. Here we evaluated the cost-effectiveness of PCV20 compared with the current standard of care (SC) and, alternatively, compared with PCV15→PPSV23, among adults aged 18 years and older in Germany. We take the payer perspective and evaluate the current German population over a lifetime.

## 2 METHODS

### 2.1 Model Description

The model utilizes a probabilistic framework and a Markov-type process to depict the lifetime risk of clinical outcomes and economic costs of pneumococcal diseases (i.e., IPD, all-cause CAP) in a hypothetical population of adults in Germany (Figure 1). The model population is initially defined based on age (i.e., in one-year increments) and risk profile (i.e., low-risk (no comorbidities considered as risk factors for pneumococcal disease), moderate-risk (at least one chronic medical condition, no immunodeficiency), or high-risk (any primary or secondary immunodeficiency) [17] for pneumococcal disease. Persons may transition to a higher risk group (i.e., from low-risk to moderate-risk, from moderate-risk to high-risk), but not to a lower risk group, during the modeling horizon. Transition probabilities between risk groups are described in the Supplement Section 2.1. Persons may receive PPSV23 alone, PCV13→PPSV23, PCV15→PPSV23, PCV20 alone, or no vaccine at model entry.

**Figure 1.**
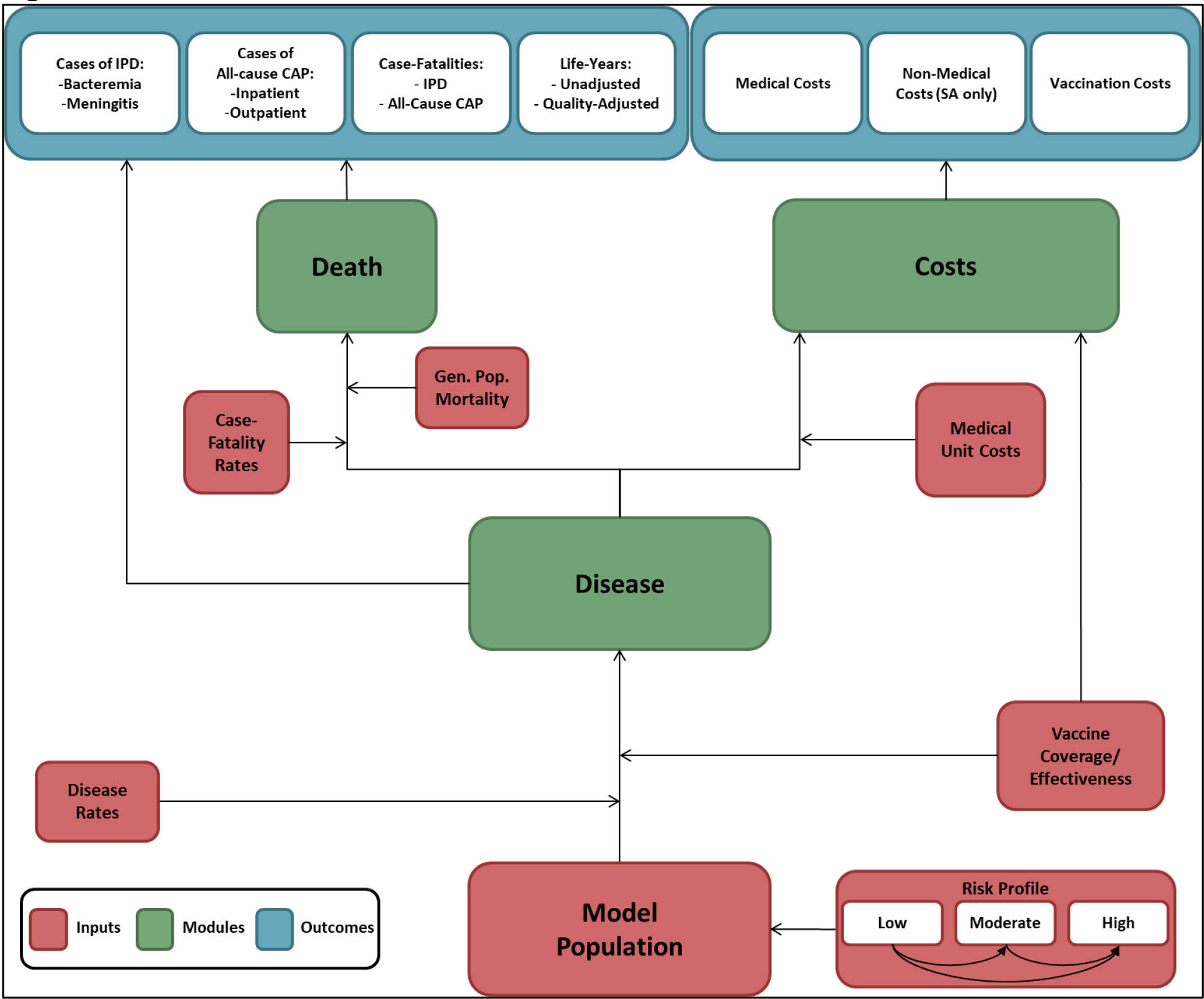
Cost-effectiveness model schematic. The figure shows the different model components. The model consists of several mutually exclusive health/disease states. Prespecified input parameters (red) determine the population and the rates at which the populations navigate through the health states. While navigating through the health states, information on the modules (disease, death, and costs) (green) are collected and transferred into outcome parameters (blue) when entering the absorbing health state “death”.

The model analyses a closed cohort that is vaccinated in year 1 with revaccination according to the vaccination strategy. Expected clinical outcomes and economic costs are projected for the model population on an annual basis, based on age, risk profile, disease/fatality risk, vaccination status, vaccine type, and time since vaccination. IPD is assumed to only include bacteremia and meningitis, and all-cause CAP is stratified by setting of care (inpatient vs. outpatient). Bacteremic CAP were classified as IPD. Persons vaccinated at model entry were at lower risk of future IPD and all-cause CAP. The magnitude of vaccine-associated risk reduction depended on the individual’s age and risk profile, clinical presentation (i.e., IPD or all-cause CAP), the vaccine(s) administered, time since vaccination, and the proportion of IPD and all-cause CAP assumed to be vaccine-preventable. Only disease caused by vaccine serotypes was reduced by vaccination.

Expected costs of medical treatment for IPD and all-cause CAP are generated based on event rates and unit costs in relation to the setting of care (i.e., inpatient vs. outpatient), age, and risk profile [18]. Costs of vaccination—including the vaccine and its administration—are tallied in the year(s) in which vaccination occurs. The value of morbidity- and mortality-related work productivity loss also is tallied in the model. Clinical outcomes and economic costs are projected over the specified period of interest (e.g., remaining years of life from model entry [max. 82 years]) for the alternative vaccination strategies considered, and include expected: numbers of cases of IPD and all-cause CAP (inpatient and outpatient); number of deaths due to IPD and all-cause CAP; number of life-years (unadjusted and quality-adjusted); costs of medical treatment for IPD and all-cause CAP; value of morbidity- and mortality-related work loss; and costs of vaccination. Future life-years and costs were discounted annually, and a healthcare system perspective was employed for the base case, while the societal perspective was assessed in sensitivity analyses. The model was programmed in Microsoft Excel 2016 as a macro-enabled workbook (.xlsm).

### 2.2 Model Estimation

Model parameter values were estimated primarily based on published literature; unpublished sources were employed as needed. Most model parameter values can be found in Table 1. Methods of estimation of model inputs are summarized below; more detailed descriptions are available in Supplementary Material.

**Table 1.**
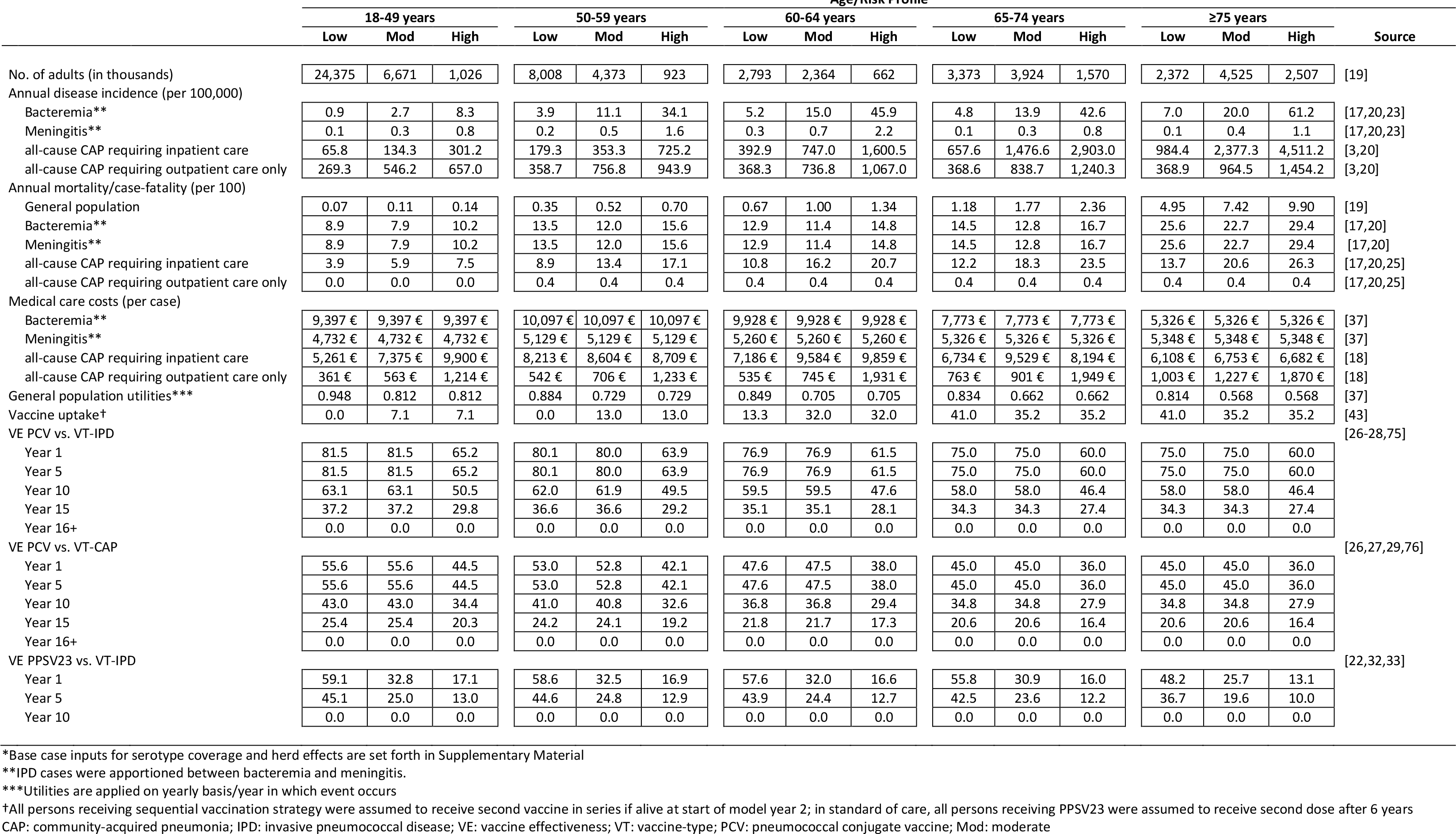
Base case input values*

#### Population

The model population included all adults in Germany aged ≥18 years (N = 69.4M), stratified across five age groups (18-49, 50-59, 60-64, 65-74, and 75-99 years) based on German census projections for 2022 [19]. The population was grouped into low-risk (immunocompetent without chronic medical conditions predisposing to pneumococcal disease), moderate-risk (immunocompetent with ≥1 chronic medical condition predisposing to pneumococcal disease), and high-risk (immunocompromised) groups based on the medical conditions specified in STIKO pneumococcal vaccination recommendations [8,17]. The prevalence of low-risk, moderate-risk and high-risk status in the German population was determined based on Pelton et al. [17] (Supplementary Material Table 1).

#### Disease Incidence

Values employed for disease incidence are summarized in Supplementary Material (Tables 4-5). Annual incidence of IPD was estimated by age and risk profile using age-specific disease rates, age-specific population distributions by risk profile, and risk-specific disease rates (for details see Supplement Section 2.3) [17,20–22]. IPD rates were apportioned between bacteremia and meningitis based on hospitalization data published by the Federal Statistical Office [23]. Age- and risk-specific annual incidence of inpatient and outpatient all-cause CAP, respectively, were based on a recent study that employed 2016-2019 data and age-specific population distributions [3,19,20].

#### Serotype Distribution and Vaccine Coverage

Values employed for serotype coverage are summarized in Supplementary Material (Tables 9-10). The proportion of IPD due to vaccine serotypes in year 1 of the modeling horizon was based on unpublished German National Reference Laboratory data provided to Pfizer [Pfizer GmbH, data on file] (Supplemental Table 9). The proportion of all-cause CAP due to pneumococcal serotypes contained in the respective vaccines [24] in year 1 of the modeling horizon was based on the prevalence of vaccine serotypes in adults admitted to hospital with all-cause CAP in Germany between 2017-2018; vaccine serotypes in pneumonia patients were detected using urine antigen detection assays (UAD) (Supplemental Table 10) [24].

#### Indirect Effects

Longstanding use of PCV13 in Germany has led to reductions in PCV13-type disease among unvaccinated adults through indirect effects (“herd effects”). We assumed PCV13-type disease has reached a steady state but incorporated potential herd effects from pediatric use of PCV15 and PCV20. The model assumes that PCV15 and PCV20 will be recommended for children in Germany by model years 2 and 3, respectively. Therefore, herd effects for serotypes unique to PCV15 were assumed to begin in model year 3, and in model year 4 for serotypes unique to PCV20 (due to later approval of PCV20 compared to PCV15 for infants). Lacking German data, reductions in the proportion of IPD and CAP due to vaccine-serotypes in adults due to indirect effects from pediatric PCV15/PCV20 vaccination were extrapolated from the reductions in IPD caused by PCV13 non-PCV7 serotypes (excluding serotype 3) observed among adults following the introduction of pediatric PCV13 in the United States (US) [Pfizer GmbH, data on file]. The relative reductions in the prevalence of PCV20 serotypes among CAP patients assumed in the model are presented in Supplemental Table 10. Serotype replacement was assumed not to occur in the base case, thus the proportion of disease due to vaccine serotypes was re-based annually (model years 3-10) to account for the reduction in overall disease [Pfizer GmbH, data on file]. No indirect effects from adult vaccination were included in the model.

#### Number of deaths and Case fatality risks (CFR)

Annual deaths were estimated by age and risk profile using age-specific disease rates and CFRs, and age-specific population distributions (Supplementary Material Tables 6-8) [17,20]. CRFs of IPD, hospitalized CAP, and outpatient CAP was based on published literature and reports [17,20,25] (Supplementary Material Table 7).Age-specific all-cause German mortality was apportioned across risk groups based on assumed relative risk for mortality and age-specific risk distributions Supplementary Material Table 8) [17,19].

#### Effectiveness of PCVs

Effectiveness data of PCVs are shown in Supplementary Material Tables 12 - 14. In year 1, effectiveness of PCVs against VT-IPD (VT-IPD) and vaccine-type CAP (VT-CAP) for low- and moderate-risk persons aged ≥18 years was based on data from the Community-Acquired Pneumonia Immunization Trial in Adults (CAPiTA) [26,27]. When discussing VT-IPD or VT-CAP cases, we mean IPD or CAP cases that are infected by serotypes covered by the vaccine a patient could have been vaccinated with in a given scenario. For high-risk persons aged 18-99 years, vaccine effectiveness (VE) against VT-IPD and VT-CAP in year 1 was assumed to be 80% of values for low/moderate-risk persons of the same age [28]. In line with findings from CAPiTA and post-hoc analyses from the trial, initial VE-PCV against VT disease was assumed to be durable for five years [26,29], then to decline 5% annually during years 6-10, 10% annually during years 11-15, and to be zero from year 16 through the end of the modelling horizon [27].

Scenario analyses were conducted using the VE assumptions used by the US Centers for Disease Control and Prevention (CDC) and Hoshi and colleagues in their evaluations of higher-valent PCVs. [30,31] For these scenarios a VE lower than base-case PCV VE was assumed, as well as steeper waning of effectiveness, lower VE against ST3 disease, lower VE in immunocompromised individuals, and higher VE against VT-CAP not caused by ST3 in low-risk adults. Details are available in Supplemental Table 24.

#### Effectiveness of PPSV23

Initial age- and risk-specific VE-PPSV23 against VT-IPD was derived from a recent real-world effectiveness study conducted in the United Kingdom aged ≥18 years [32] (Supplemental Table 13). VE-PPSV23 against VT-IPD was assumed to decline linearly beyond the first year of the modelling horizon, to 76.2% of initial VE by year five and to no effectiveness by year ten [33]. Consistent with evidence from recent publications, VE- PPSV23 against VT-CAP was assumed to be 0% [34–36]. No boosting or blunting of immune response for the serotypes shared between both vaccines was assumed from vaccination with both PCV and PPSV23; the individual was assumed to benefit from both vaccines with the effects of each vaccine waning independently. Scenario analyses used CDC assumptions on PPSV23 VE (Supplemental Table 24). The initial VE of PPSV23 assumed by CDC was higher than our base case estimate, and the waning of VE was longer (15-year waning period). CDC also assumed some effectiveness against VT-CAP in contrast to the base case assumption used in previous assessments, while serotype 3 VE was assumed to be the same as the overall VE of PPSV23. Details are available in Supplemental Table 23.

#### Costs

Age-specific payer costs of IPD hospitalization (bacteremia and meningitis, respectively) were from a previously published cost-effectiveness analysis of pneumococcal vaccination in Germany which employed estimates based on national tariffs/diagnosis-related groups [37,38]. The costs were updated to the calendar year 2022 and expressed in Euro (2022 Euro) using the healthcare component of the consumer price index [37,38]. Age- and risk-specific costs of all-cause CAP requiring hospitalization and outpatient care only, respectively, were derived healthcare claims data and included insurer paid amounts for all healthcare encounters that occurred within 30 days of the start of the CAP episode (i.e., date of hospital admission or date of outpatient encounter for CAP) [18].

The prices of PPSV23 (33.88 €), PCV13 (72.50 €), PCV15 (76.74 €), and PCV20 (76.74 €) were based on published list prices [39]. The cost of vaccine administration was assumed to be 8.19 € [37] (Supplementary Material Tables 16-17).

#### Utilities

General population health state utilities were the same as those employed in the aforementioned published cost-effectiveness analysis (Supplementary Material Table 15) [37]. An annual utility decrement for persons who experienced IPD or CAP was applied during the year in which the event occurred. The one-year excess QALY loss attributed to all-cause CAP or IPD was 0.13 [40] and to outpatient all-cause CAP was 0.004 [27,41].

### 2.3 Analyses

#### Base Case

Clinical outcomes and economic costs were projected over a lifetime (82 years) for the model population under PCV20 and SC (i.e., age/risk-dependent use of PPSV23 or PCV13→PPSV23 with PPSV23 revaccination). Vaccine coverage for PCV20 and PCV15-◊PPSV23 was based on published estimates of vaccine coverage for adults aged 18-59 with underlying conditions and all adults aged ≥60 years in 2020 and was assumed to be the same regardless of vaccine strategy (Supplementary Material Tables 18-19) [42,43]. In SC, all persons who received PCV13 at model entry were assumed to receive PPSV23 if alive in year 2; all persons who received a dose of PPSV23 (i.e., at model entry or in year 2) as part of SC were also assumed to receive a second dose of PPSV23 if alive 6 years after administration of the first dose. To convert values (costs and life years) received in the future to current time equivalents, we discounted future costs, life-years (LYs), and quality-adjusted LYs (QALYs) at 3.0% annually; analyses were conducted from the payer perspective (i.e., excluding patient paid amounts) [44,45].

#### Sensitivity

Sensitivity analyses were conducted to evaluate the robustness of base case results to changes in key model parameter values, including: changes to incidence parameters, case fatality rates, costs, proportion of all-cause CAP requiring hospitalization, proportion of PPSV23 revaccination (SC only), vaccine effectiveness, utilities, and herd effects [33,46]. For these parameters, we changed the single parameters.

#### Scenario

Several scenario analyses were conducted in which more than one parameter was varied and the impact on the results was tested (Supplementary Material Section 5). First, PCV20 and PCV15→PPSV23 (without PPSV23 revaccination) were compared; second, the societal perspective was adopted; third, the set of vaccine effectiveness parameters (including waning) were changed using sources from independent bodies such as CDC; forth, several different sources for case fatality rates and costs were changed; and finally, a scenario was analyzed where the lowest PCV and high PPV efficacy assumptions and low CAP cost assumptions were combined.

## 3 RESULTS

Throughout a lifetime, use of PCV20 in lieu of SC in the German adult population would prevent an additional 2,801 (1.3%) IPD cases, 51,532 (0.2%) hospitalized CAP cases, 26,368 (0.2%) outpatient CAP cases, and 10,946 (0.2%) disease-related deaths (Table 2). This translates to 107 prevented IPD cases, 2,038 hospitalized CAP cases, and 1,048 outpatient CAP cases per year. Throughout a lifetime cohort of German adults, PCV20 vaccination would save 363.2 million € of the medical care costs and vaccination costs would increase by 6.5 million €, resulting in a 356.7 million € reduction in total costs. Also, PCV20 vaccination would save 74,694 additional total LYs, and 49,655 additional QALYs. The PCV20 strategy is dominant compared to SC (i.e., it resulted in more life years and fewer costs). In one-way sensitivity analyses assuming lower pneumococcal disease incidence, lower case fatality risks, treatment costs, lower PCV20 VE, or higher VE of PPSV23, PCV20 remained cost-saving (Table 2).

**Table 2.** Clinical and economic outcomes of PCV20 vs PPSV23/PCV13+PPSV23 among adults ages 18-99 in Germany (N=69,466,087)

| A. Base Case analysis | PPSV23/PCV13+PPSV23<br>(Standard of Care) | PCV20 | Difference |
| --- | --- | --- | --- |
| **Clinical Outcomes** |  |  |  |
| No. of Cases |  |  |  |
| IPD | 216,227 | 213,426 | -2,801 |
| ACP |  |  |  |
| Hospitalized | 22,776,294 | 22,724,762 | -51,532 |
| Requiring Outpatient Care Only | 13,088,903 | 13,062,535 | -26,368 |
| No. of Deaths | 4,606,187 | 4,595,242 | -10,946 |
| Total Life-Years (discounted) | 1,287,861,006 | 1,287,935,699 | 74,694 |
| Total Quality-Adjusted Life-Years (discounted) | 1,013,729,588 | 1,013,779,243 | 49,655 |
| **Economic Outcomes** |  |  |  |
| Total Costs (millions) |  |  |  |
| Medical Care | 89,989.62 € | 89,626.40 € | -363.22 € |
| Vaccination | 767.13 € | 773.65 € | 6.52 € |
| Medical Care + Vaccination | 90,756.75 € | 90,400.05 € | -356.70 € |
| Non-medical Costs | 119,913.00 € | 119,589.00 € | -324.00 € |
| Total Costs | 210,669.75 € | 209,989.05 € | -680.70 € |
| Cost-Effectiveness (PCV20 vs PPSV23/PCV13+PPSV23)) |  |  |  |
| Cost per LY | --- | --- | Dominant |
| Cost per QALY | --- | --- | Dominant |
| **B. Sensitivity analyses** |  |  |  |
|  | Difference<br>in Costs (millions) | Difference<br>in LYs | Incremental<br>Cost per LY |
| Base Case | -356.70 € | 74,694 | Dominant |
| Bacteremia Incidence (-25%) | -353.10 € | 73,801 | Dominant |
| Bacteremia Incidence (+25%) | -360.30 € | 75,586 | Dominant |
| Meningitis Incidence (-25%) | -356.63 € | 74,670 | Dominant |
| Meningitis Incidence (+25%) | -356.77 € | 74,717 | Dominant |
| Inpatient Pneumonia Incidence (-25%) | -281.22 € | 57,903 | Dominant |
| Inpatient Pneumonia Incidence (+25%) | -430.16 € | 91,063 | Dominant |
| Outpatient Pneumonia Incidence (-25%) | -350.40 € | 74,486 | Dominant |
| Outpatient Pneumonia Incidence (+25%) | -363.00 € | 74,901 | Dominant |
| Bacteremia Case-Fatality Risks (-25%) | -356.94 € | 73,801 | Dominant |
| Bacteremia Case-Fatality Risks (+25%) | -356.46 € | 75,586 | Dominant |
| Meningitis Case-Fatality Risks (-25%) | -356.71 € | 74,670 | Dominant |
| Meningitis Case-Fatality Risks (+25%) | -356.69 € | 74,717 | Dominant |
| Inpatient Pneumonia Case-Fatality Risks (-25%) | -362.84 € | 57,903 | Dominant |
| Inpatient Pneumonia Case-Fatality Risks (+25%) | -350.69 € | 91,063 | Dominant |
| Outpatient Pneumonia Case-Fatality Risks (-25%) | -356.76 € | 74,486 | Dominant |
| Outpatient Pneumonia Case-Fatality Risks (+25%) | -356.64 € | 74,901 | Dominant |
| Bacteremia Costs (-25%) | -352.86 € | 74,694 | Dominant |
| Bacteremia Costs (+25%) | -360.54 € | 74,694 | Dominant |
| Meningitis Costs (-25%) | -356.62 € | 74,694 | Dominant |
| Meningitis Costs (+25%) | -356.78 € | 74,694 | Dominant |
| Inpatient Pneumonia Costs (-25%) | -276.17 € | 74,694 | Dominant |
| Inpatient Pneumonia Costs (+25%) | -437.23 € | 74,694 | Dominant |
| Outpatient Pneumonia Costs (-25%) | -350.34 € | 74,694 | Dominant |
| Outpatient Pneumonia Costs (+25%) | -363.06 € | 74,694 | Dominant |
| Costs (-25%) | -265.90 € | 74,694 | Dominant |
| Costs (+25%) | -447.51 € | 74,694 | Dominant |
| PPSV23 VE-IPD (-25%) | -359.47 € | 75,385 | Dominant |
| PPSV23 VE-IPD (+25%) | -353.93 € | 74,002 | Dominant |
| VE-PPSV23 vs. VT-CAP from Suzuki et al. [33] | -248.25 € | 52,534 | Dominant |
| VE-PPSV23 vs. VT-CAP from Lawrence et al. [46] | -291.88 € | 60,599 | Dominant |
| VE-PPSV23 vs. VT-CAP Lawrence w/ 10-yr waning [46] | -252.86 € | 52,329 | Dominant |
| PCV13/PCV20 VE-IPD (-25%) | -350.20 € | 73,075 | Dominant |
| PCV13/PCV20 VE-IPD (+25%) | -363.18 € | 76,308 | Dominant |
| PCV13/PCV20 VE-CAP (-25%) | -269.53 € | 56,933 | Dominant |
| PCV13/PCV20 VE-CAP (+25%) | -443.90 € | 92,462 | Dominant |
| No Herd Effects | -472.74 € | 98,567 | Dominant |
| Full Serotype Replacement for Herd Effects | -353.95 € | 74,046 | Dominant |
| 10% Revaccination with PPSV23 (SC Only) | -472.74 € | 98,567 | Dominant |

PCV: pneumococcal conjugate vaccines; PCV20: 20-valent PCV; PCV13: 13-valent PCV; PPSV23: 23-valent pneumococcal polysaccharide vaccine; CAP: all-cause community-acquired pneumonia; IPD: invasive pneumococcal disease; LY: life year; QALY: quality adjusted life year; VE: vaccine effectiveness; VT: vaccine-type; SC: standard of care

The impact of model assumption was further explored in several scenarios varying more than one model parameter (Table 3). In one scenario PCV20 was compared to PCV15→PPSV23 instead of SC (Table 3, scenario 1). In this scenario PCV20 continued to result in better outcomes and would prevent 928 cases of IPD, 12,485 cases of CAP requiring hospitalization, 5,978 cases of CAP in the outpatient setting, and 2,776 disease-related deaths more than PCV15→PPSV23. In scenario 1, medical care and vaccination costs would be 97.0 million € and 357.0 million € lower, respectively, and total costs would be lower by 454.0 million €. PCV20 would add 20,277 total LYs and 13,298 QALYs and therefore would be also cost-saving compared to PCV15→PPSV23.

**Table 3.** Clinical and economic outcomes -- scenario analyses

| Scenario (Supplemental Material Section 5) |  | Difference in Clinical Outcomes |  |  |  |  |  | Difference in Economic Outcomes (disc) |  |  | Cost-Effectiveness (disc) |  |
| --- | --- | --- | --- | --- | --- | --- | --- | --- | --- | --- | --- | --- |
|  |  | IPD | CAP inp | CAP outp | No. of Deaths | Total LY (disc) | Total QALY (disc) | Medical Care Cost (millions) | Vaccination Cost (millions) | Total Cost (millions) | Cost per LY | Cost per QALY |
|  | Base Case | -2,801 | -51,532 | -26,368 | -10,946 | 74,694 | 49,655 | -363.22 € | 6.52 € | -356.70 € | Dominant | Dominant |
| 1 | PCV20 vs PCV15+PPSV23 | -928 | -12,485 | -5,978 | -2,776 | 20,277 | 13,398 | -97.02 € | -357.03 € | -454.04 € | Dominant | Dominant |
| 2 | Societal perspective | -2,801 | -51,532 | -26,368 | -10,946 | 74,694 | 49,655 | -363.22 € | 6.52 € | -680.70 € | Dominant | Dominant |
| 3 | Lower VE for PCV13, higher VE for PPSV23 [31,77] | 947 | -644 | -1,460 | 341 | 3,664 | 3,169 | -17.08 € | 6.51 € | -10.57 € | Dominant | Dominant |
| 4 | CFR for CAP reported from Saxony, IPD CFR reported from UK [22,78] | -2,767 | -50,225 | -25,874 | -14,454 | 95,536 | 61,301 | -356.90 € | 8.24 € | -348.67 € | Dominant | Dominant |
| 5 | CFR for CAP and CAP used in RKI 2016 CE-model [25] | -2,834 | -52,899 | -26,956 | -8,080 | 51,348 | 36,274 | -370.33 € | 4.77 € | -365.56 € | Dominant | Dominant |
| 6 | Lower medical costs as used for RKI 2016 CE-model [25] | -2,801 | -51,532 | -26,368 | -10,946 | 74,694 | 49,655 | -181.91 € | 6.52 € | -175.39 € | Dominant | Dominant |
| 7 | Lower VE for PCV13, higher VE for PPSV23 and lower medical cost as used for RKI 2016 CE-model [25,31,77] | 947 | -644 | -1,460 | 341 | 3,664 | 3,169 | -5.73 € | 6.51 € | 0.78 € | 214 € | 247 € |

IPD: invasive pneumococcal disease; CAP: community-acquired pneumonia; inp: inpatient; outp: outpatient; disc: discounted; LY: life-year; QALY: quality adjusted life year; CDC: US Centers for Disease Control and Prevention; CRF: Case Fatality Risk; RKI Robert Koch Institut

Other scenario analyses evaluated the effects of model inputs assuming a lower initial VE and faster wanting pattern for PCV13 and PCV20, reduced PCV13/PCV20 VE against serotype 3, combined with full effectiveness of PPSV23 against serotype 3, and some effectiveness of PPSV23 against CAP (Table 3, scenario 3, for model inputs see Supplemental Table 24). This scenario modelled smaller clinical benefits and smaller cost savings of PCV20 compared to SC and PCV20 would only prevent 947 cases of IPD, 644 cases of hospitalized CAP, and 1,460 cases of CAP in the outpatient setting more than the SC. Medical care would be lower by 17.1 million € and vaccination costs would increase by 6.5 million €. Thus, total costs would be lower by 10.6 million €. PCV20 would add 3,664 total LYs and 3,169 QALYs and was therefore dominant compared with SC. Scenario analyses assuming a societal perspective instead of a payer’s perspective, alternative CFRs, and alternative costs did not substantially change the total LY and QALY saved, or the cost saved compared to the base case and PCV20 remained cost-saving (Table 3, scenarios 2, 4, 5, and 6).

In only in the most extreme scenario that assumed reduced VE of PCV20 against serotype 3, combined with full effectiveness of PPSV23 against serotype 3, some effectiveness of PPSV23 against CAP as well as lower treatment costs for pneumococcal disease PCV20 was not cost saving, but showed a cost-effectiveness ratio of 214 € per life year and 247 € per QALY saved (Table 3, scenario 7).

## 4 DISCUSSION

To the best of our knowledge, this is the first evaluation of the potential public health and economic impacts of next generation pneumococcal vaccination strategies for adults in Germany. The results of our analyses suggest that replacing SC with a single dose of PCV20 for adults aged ≥60 years without underlying diseases and adults aged 18-59 years with moderate- and high-risk conditions would reduce the burden of pneumococcal disease and pneumococcal-related deaths, thereby reducing health care costs and resource usage and would be cost saving overall. Findings also suggest PCV20 would prevent more cases of disease and associated deaths and would reduce total costs compared with PCV15 followed by [47–51]PPSV23. PCV20 remained cost-saving in one-way sensitivity analyses and scenario analyses that varied inputs for cost, disease burden, and vaccine effectiveness.

Our findings are consistent with several recently published analyses considering use of PCV20 in other settings, including England, Denmark, the US, and Canada [52–54]. Mendes and colleagues and Olsen and colleagues found that PCV20 was cost saving compared with current recommendations (i.e., PPSV23) in a cohort of all adults aged ≥65 years and moderate- and high-risk adults aged 18-64 years in England and Denmark, respectively. [52,53] In an analysis conducted by the US CDC Advisory Committee on Immunization Practices (ACIP) considering all adults aged ≥65 years and adults aged 19-64 years with chronic medical or immunocompromising conditions, a single dose of PCV20 was found to be cost saving compared with 2019 ACIP recommendations, which included age/risk dependent use of [31] PPSV23 alone or PCV13→PPSV23 [31]. Likewise, recent analyses by the Public Health Agency of Canada found PCV20 was likely to be cost-effective in Canadian adults aged ≥65 years [55]. We note that in a recent analysis by Smith et al., PCV20 use (vs. 2019 ACIP recommendations) among elderly US adults was found to cost $172,491 per QALY, however, differences in findings (compared with aforementioned US CDC analysis) are believed to be due, at least in part, to variation in model populations (e.g., Smith et al. excluded immunocompromised patients) as well as key parameter values (e.g., rates of pneumonia were assumed by Smith et al. to be lower) [54]. Comparisons between the present study and the Smith et al study are complicated by the differences between the German and US health systems, the standard of care comparator vaccine, as well as difference in model structure and populations.

However, a key difference appears to be the incidence of vaccine-preventable disease in comparable population segments; with higher incidence PCV20-type CAP among unvaccinated low-risk adults 65 years of age assumed in our model than in the model by Smith et al.. A similar pattern is seen across age and risk segments included in the models, and the present study also including high-risk adults amongst whom the vaccine is more likely to be cost-effective given their elevated rates of disease, a group excluded from the US analysis.

Our analyses have several limitations. First, we aimed at minimizing a potential conflict of interest arising from the author affiliation by strictly adhering to published practice guidelines [44,45,56–60], using the most recent and highest quality data, providing detailed description of the methodology, and by addressing uncertainties with numerous sensitivity analyses. Specifically, we addressed uncertainties around vaccine effectiveness and duration of protection by performing sensitivity analyses that use model inputs for PCV20 and PPSV23 effectiveness based on assumptions by the US CDC [30] and cost data used for the RKI model that were higher for meningitis, similar for bacteremia, and lower for CAP when compared to the respective medical costs in the base case. [25]. Second, we aimed at using model parameters based on German data. However, this was not always possible. When German data were not available, data from other high-income European countries with similar health care systems were applied. It was assumed that reductions in disease attributable to the seven new serotypes included in PCV20 (i.e., PCV20 non-PCV13 serotypes) among adults via pediatric use of next generation vaccines higher-valent PCVs would be similar to reductions observed in PCV13 non-PCV7 type disease among adults following the introduction of pediatric PCV13 and that serotype replacement would not occur; however, the extent to which the magnitude and patterns of vaccine-type disease reduction may vary and whether replacement will occur is unknown.

In the base case, we did not model serotype replacement, explicitly. It is unknown to what extent replacement of serotypes newly covered by PCV15 and PCV20 will take place.

Model simulations suggest that replacement may be less for high-valency PCVs [61,62]. However, we tested the impact of our base case assumptions on the overall results. The analyses were not very sensitive to assumptions on serotype replacement.

Also, the model did not consider adverse events originating from vaccination by PCV20, PCV15, PPSV23 or PCV13. As adverse events after pneumococcal vaccination resulting in healthcare seeking are rare and event rates are similar for the different pneumococcal vaccines, it is unlikely that the omission of adverse events from our model has introduced bias that would have affected our main conclusions. On the contrary, PCV20 is only administered once. Hence, the risk of adverse events is lower in the strategy where PCV20 is administered once compared to other strategies with vaccination with comparable safety profile administered more than once.

Another area of uncertainty surrounds vaccine effectiveness. Next generation PCVs have been licensed based on studies demonstrating non-inferior immunogenicity compared the PCV13 or PPV13 [14,63–65]. Effectiveness of PCV20 and PCV15, respectively, against VT-IPD and VT-CAP was therefore assumed to be the same as VE-PCV13 against VT disease from CAPiTA [26]. While it is anticipated that the additional serotypes in higher-valent PCVs will perform similarly to PCV13 in terms of disease prevention, it is possible that the effectiveness of PCV20 and/or PCV15 may [32]differ from PCV13. Masala and colleagues estimated that increased valency may decrease vaccine effects [62]. We tested this by simulating lower effectiveness of PCVs. Changes to the assumptions resulted in fewer cases avoided and smaller life expectancy increase. However, PCV20 kept being the dominant strategy avoiding more cases, increasing life-expectancy while costing less than the standard of care. The effectiveness of PPSV23 against VT-IPD was assumed to decline linearly as observed in a test-negative design study from Japan [33]. We also note that the findings presented herein likely underestimate the impact of PCV20 use among adults in Germany because multiple studies evaluating the impact of PCV13 against all-cause CAP have reported larger reductions than what would be expected if accounting only for etiologically confirmed vaccine-type pneumonia [66–68].

Some recently published real-world data analyses suggest that PPSV23 may provide limited protection against VT-CAP, but because there is considerable inconsistency across studies, we do not believe these data are sufficiently robust to employ in base case analyses [33,34,36,46,69]. Nonetheless, we conducted scenario analyses that assumed moderate PPSV23 effectiveness against VT-CAP based on findings from Lawrence et al. and Suzuki et al., respectively [33,46]. Under these assumptions, SC and PCV15→PPSV23 prevented more cases of all-cause CAP and associated deaths (compared with base case analyses), but PCV20 remained dominant against both strategies in all analyses conducted, with cost savings ranging from 248-292 million € (vs. SC) and from 432-442 million € (vs. PCV15→PPSV23).

Current STIKO recommendations include revaccination with PPSV23 on a case-by-case basis only, however, estimates of the proportion of adults who are revaccinated are not available. We therefore assumed in base case analyses that all persons in the SC strategy who received PPSV23 alone or PCV13→PPSV23 at model entry received a second dose of PPSV23 (i.e., revaccination) 6 years later. Because this likely overestimates the cost—and associated benefits—of vaccination in the SC strategy, we conducted a sensitivity analysis in which only 10% of persons were revaccinated. With lower rates of revaccination, the costs of vaccination in SC dropped dramatically (nearly 50% compared with base case). Nonetheless, total costs with PCV20 were still slightly lower and PCV20 remained dominant against SC.

We used case fatality rates from a recently published article on the burden of pneumococcal disease [20]. The mortality rates of CAP estimated in that paper were higher than previous estimates (2015) [25], so we tested the impact of more recent higher CFR in sensitivity analyses using lower CFR, which were applied by a previous model of PCV13 cost-effectiveness [25]. PCV20 strategy was still the dominating strategy, meaning it saved more life-years while costing less than the standard of care strategy.

The analyses presented herein were conducted from the payer perspective which does not reflect the true costs of medical care, such as patient paid amounts. Finally, our model, like all such health economic models, simplifies reality. For example, long-term consequences of pneumococcal disease—including exacerbations of underlying conditions such as COPD or asthma—also were not considered, despite evidence that such sequelae are common and costly [70–74].

## 5 CONCLUSIONS

Use of a single dose of PCV20 among adults aged ≥60 years and adults aged 18-59 years with moderate- and high-risk conditions would reduce the burden of pneumococcal diseases, save more lives, and would be cost saving compared with SC. PCV20 alone would also be cost saving compared with use of PCV15→PPSV23 [47–51]. The availability of a single dose PCV20 strategy will likely improve vaccine uptake compared to sequential regimens which are often not completed, leading to greater benefit in terms of pneumococcal disease prevention. The simplicity of a single dose strategy for pneumococcal disease prevention among adults may also reduce burden on providers, payers, and the healthcare system overall.

Future research examining the real-world impact of PCV20 use on pneumococcal disease burden in Germany should aim to capture other potential benefits of a single dose vaccination strategy for pneumococcal disease prevention among adults.

## Data Availability

All data produced in the present study are available upon reasonable request to the authors

## Acknowledgement

The authors would like to thank Reiko Sato and Julia Schiffner-Rohe for many valuable discussions and Mark van der Linden for providing IPD data. Further, we would like to thank Ahuva Hanau for medical writing.

