## Supplemental for "Cost-effectiveness of use of 20-valent pneumococcal conjugate vaccine among adults in Germany"

Supplementary Material: Methods

### 1 Model Structure

A probabilistic model with a Markov-type process was used to depict the lifetime risk of clinical outcomes and economic costs of pneumococcal disease (i.e., invasive pneumococcal disease [IPD], all-cause community-acquired pneumonia [all-cause CAP]) in a hypothetical population of German adults (Figure 1). The model has been employed in previously published analyses comparing use of PCV20 with various vaccination strategies in England [1]. The model population is initially characterized based on age (i.e., in one-year increments) and risk profile (i.e., high-risk [immunocompromised], moderate-risk [immunocompetent with ≥1 underlying medical condition], or low-risk [immunocompetent without underlying medical conditions]). Persons may transition to a higher risk group (i.e., from low-risk to moderate-risk, from moderate-risk to high-risk), but not to a lower risk group, during the modeling horizon. Persons in the model population may be assumed to receive PPSV23 alone, PCV13→PPSV23, PCV20 alone, PCV15→PPSV23, or no vaccine at model entry; persons who received PPSV23 alone or PCV13→PPSV23 may be assumed to be revaccinated with PPSV23 6 years after receiving their initial dose.

Expected clinical outcomes and economic costs are projected for the model population on an annual basis, based on age, risk profile, disease/fatality risks, vaccination status, vaccine type, and time since vaccination. IPD is assumed to include bacteremia and meningitis, and all-cause CAP is stratified by care setting (inpatient vs. outpatient). Persons vaccinated at model entry may be at lower risk of future IPD and all-cause CAP; the magnitude of vaccine-associated risk reduction depends on clinical presentation (i.e., IPD or all-cause CAP), as well as the vaccine(s) received, age, time since vaccination, and risk profile. Risk of death from IPD, all-cause CAP requiring inpatient care, and other causes (i.e., other than IPD and all-cause CAP) depends upon age and risk profile.

Expected costs of medical treatment for IPD and all-cause CAP are generated based on event rates and unit costs in relation to the setting of care (i.e., inpatient vs. outpatient), age, and risk profile. Costs of vaccination—including the vaccine and its administration—are tallied in the year(s) in which the vaccination occurs (e.g., year 1 for single vaccine strategies, years 1 and 2 for sequential vaccination). The value of morbidity- and mortality-related work loss also is tallied in the model.

Clinical outcomes and economic costs are projected over the modeling horizon for the alternative vaccination strategies considered and include expected:

- Numbers of cases of IPD and all-cause CAP (inpatient and outpatient)
- Number of deaths due to IPD and all-cause CAP
- Number of life-years (unadjusted and quality-adjusted)
- Costs of medical treatment for IPD and all-cause CAP
- Costs of vaccination
- Value of morbidity- and mortality-related work loss.

The cost-effectiveness of alternative vaccine strategies is calculated in terms of incremental cost per life-year (LY) gained and per quality-adjusted life-year (QALY) gained. Future life-years and costs may be discounted annually, and a healthcare system or societal perspective may be employed.

**Figure 1. Cost-effectiveness model schematic**

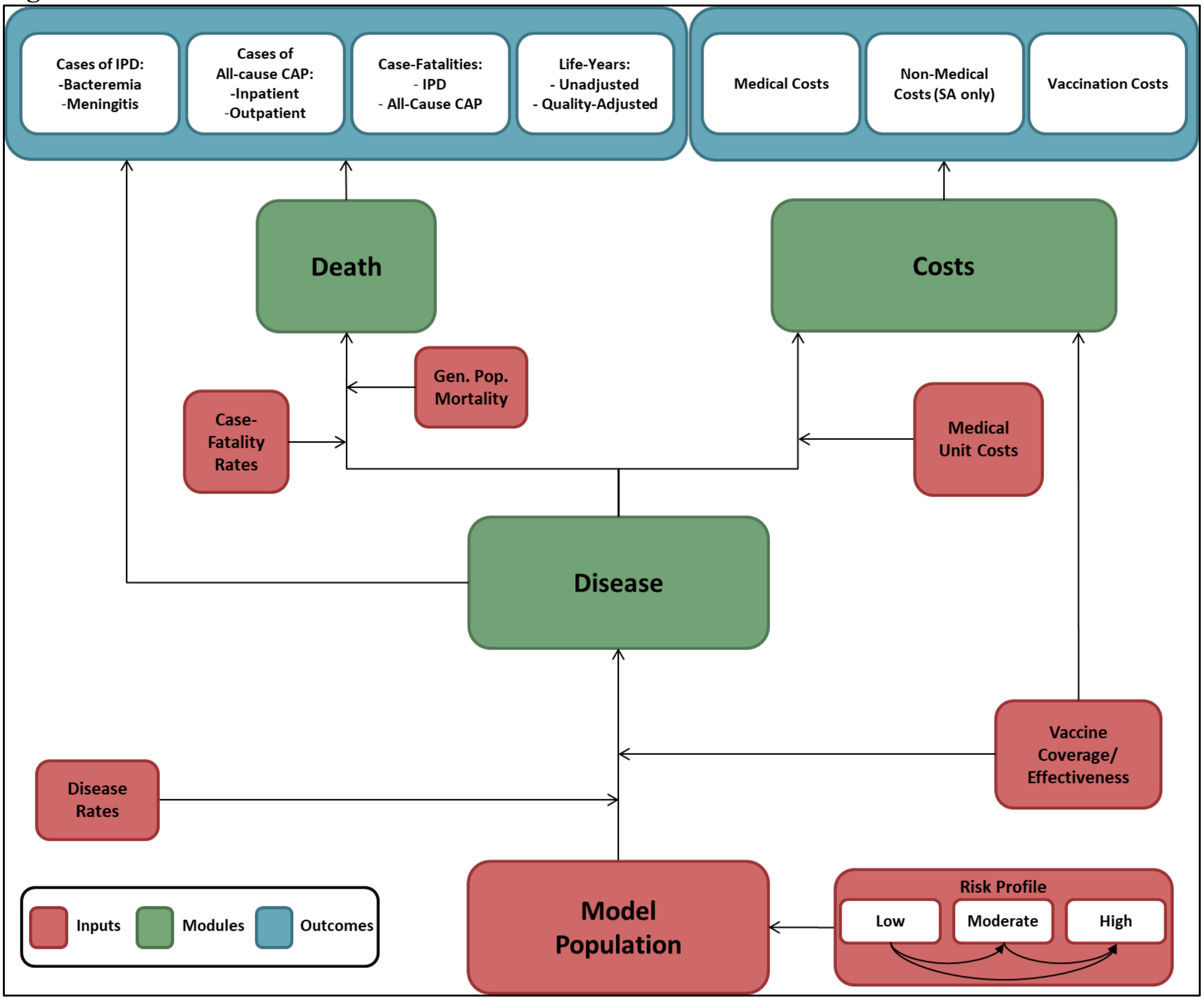


#### 2 Estimation of Model Inputs for Base Case Analyses

##### 2.1 Model Population

The model population included—at model entry—low-risk (N=40.9M), moderate-risk (N=21.9M), and high-risk (N=6.7M) persons aged 18-99 years. Estimates of the size of the German population aged ≥18 years, in one-year increments, were obtained from German census projections for 2022 [2].

Risk groups were defined based on the conditions included in STIKO pneumococcal vaccination recommendations and unpublished data generated as part of a published analysis[3,4], as follows:

- Low risk: immunocompetent without chronic medical conditions (CMCs)
- Moderate Risk: immunocompetent with ≥1 CMC; and
- High-risk: immunocompromised.

**Table 1. Distribution of model population by age and risk profile based on Pelton et al. [3]**

| Age Group (years) | Risk Profile | | |
|---|---|---|---|
| | Low | Moderate | High |
| 18-49 | 76.0% | 20.8% | 3.2% |
| 50-59 | 60.2% | 32.9% | 6.9% |
| 60-64 | 48.0% | 40.6% | 11.4% |
| 65-74 | 38.0% | 44.3% | 17.7% |
| 75-99 | 25.2% | 48.1% | 26.7% |

Transition probabilities between risk groups were determined by assigning the midpoint age within each age group in the model (e.g., 18-49, 50-59, etc.) the distribution of low, moderate, and high risk as specified in the model inputs, and the distribution of risk in each single year of age was determined by linear interpolation and extrapolation from those midpoint ages. The risk transition probabilities were then calculated based on those single-year percentages of persons at low, moderate, or high risk at the end of the year after accounting for mortality, so that the risk distributions were consistent across each single-year cohort at the same age.

#### 2.2 Vaccine Coverage

The standard of care strategy involved use of PPSV23 alone at model entry among persons aged 18-59 years who are immunocompetent with underlying chronic medical conditions (i.e., moderate-risk) and among persons aged 60-99 years who are immunocompetent with and without underlying chronic medical conditions (i.e., low- and moderate-risk), and PCV13→PPSV23 among immunocompromised (i.e., high-risk) persons aged 60-99 years. Vaccine coverage was based on published estimates of 2022 vaccine coverage in the season 21/22 for adults aged 18-59 with underlying conditions and all adults aged ≥60 years [5,6]. All persons who received PCV13 at model entry were assumed to receive PPSV23 if alive in year 2; all persons who received a dose of PPSV23 (i.e., at model entry or in year 2) were also assumed to receive a second dose of PPSV23 if alive 6 years after administration of the first dose.

**Table 2. Vaccine coverage with SC**

| Age Group (years) | Risk Profile | | |
|---|---|---|---|
| | Low | Moderate | High |
| 18-49 | No vaccine: 100.0% | PPSV23: 7.1% | PCV13→PPSV23: 7.1% |
| 50-59 | No vaccine: 100.0% | PPSV23: 13% | PCV13→PPSV23: 13% |
| 60-64 | PPSV23: 13.3% | PPSV23: 32% | PCV13→PPSV23: 32% |
| 65-74 | PPSV23: 41% | PPSV23: 35.2% | PCV13→PPSV23: 35.2% |
| 75-99 | PPSV23: 41% | PPSV23: 35.2% | PCV13→PPSV23: 35.2% |

The hypothetical strategy involved use of PCV20 at model entry (i.e., in lieu of SC) among persons aged 18-59 years who are immunocompetent with underlying chronic medical conditions or who are immunocompromised (i.e., moderate- and high-risk) and among all persons (i.e., low-, moderate-, and high-risk) aged 60-99 years. Vaccine coverage was assumed to be the same as that for corresponding age/risk groups in SC.

**Table 3. Vaccine coverage with PCV20**

| Age Group (years) | Risk Profile | | |
|---|---|---|---|
| | Low | Moderate | High |
| 18-49 | 0% | 7.1% | 7.1% |
| 50-59 | 0% | 13% | 13% |
| 60-64 | 13.3% | 32% | 32% |
| 65-74 | 41% | 35.2% | 35.2% |
| 75-99 | 41% | 35.2% | 35.2% |

#### 2.3 Disease Rates

**IPD.** Annual rates of IPD were estimated by age group and risk profile using age-specific disease rates (adjusted for laboratory and hospital under-reporting) from Deb et al. [7], age-specific population distributions by risk profile[3], and risk-specific disease rates for IPD from Deb et al. [7]. IPD were apportioned between bacteremia and meningitis based on hospitalization data published by the Federal Statistical Office; the proportion of IPD attributable to bacteremia was assumed to be 91.1% for persons aged 18-49 years, 95.4% for persons aged 50-64 years, and 98.2% for persons aged 65-99 years [8]. Assumed indirect effects from future use of PCV15 and PCV20 in children on disease serotype distribution and overall disease incidence is described in Section 2.6.

**Table 4. Annual incidence rates of IPD (per 100,000) in model year 1**

| Age Group (years) | Bacteremia | | | Meningitis | | |
|---|---|---|---|---|---|---|
| | Risk Profile | | | Risk Profile | | |
| | Low | Moderate | High | Low | Moderate | High |
| 18-49 | 0.9 | 2.7 | 8.3 | 0.1 | 0.3 | 0.8 |
| 50-59 | 3.9 | 11.1 | 34.1 | 0.2 | 0.5 | 1.6 |
| 60-64 | 5.2 | 15.0 | 45.9 | 0.3 | 0.7 | 2.2 |
| 65-74 | 4.8 | 13.9 | 42.6 | 0.1 | 0.3 | 0.8 |
| 75-99 | 7.0 | 20.0 | 61.2 | 0.1 | 0.4 | 1.1 |

**All-Cause CAP.** Age- and risk-specific annual rates of inpatient and outpatient all-cause CAP, respectively, were based on a recent study that estimated CAP rates among adults in Germany aged ≥16 years who were diagnosed with incident CAP in 2016-2019 (based on evidence of ICD-10 codes plus other inclusion criteria) [7,9]. Assumed indirect effects from future use of PCV15 and PCV20 in children on disease serotype distribution and overall incidence of all-cause CAP requiring inpatient care and outpatient care only, respectively, is described in Section 2.6.

**Table 5. Annual incidence rates of all-cause CAP (per 100,000) in model year 1**

| Age Group (years) | Inpatient Care | | | Outpatient Care Only | | |
|---|---|---|---|---|---|---|
| | Risk Profile | | | Risk Profile | | |
| | Low | Moderate | High | Low | Moderate | High |
| 18-49 | 65.8 | 134.3 | 301.2 | 269.3 | 546.2 | 657.0 |
| 50-59 | 179.3 | 353.3 | 725.2 | 358.7 | 756.8 | 943.9 |
| 60-64 | 392.9 | 747.0 | 1,600.5 | 368.3 | 736.8 | 1,067.0 |
| 65-74 | 657.6 | 1,476.6 | 2,903.0 | 368.6 | 838.7 | 1,240.3 |
| 75-99 | 984.4 | 2,377.3 | 4,511.2 | 368.9 | 964.5 | 1,454.2 |

#### 2.4 Case-Fatality Risks

**IPD.** Age -specific IPD case-fatality risks (CFRs) [7], age-specific population distributions by risk profile [3], and risk ratios[7] CFRs were assumed to be the same for bacteremia and meningitis.

**Table 6. Case-fatality risks for IPD (per 100)**

| Age Group (years) | Bacteremia | | | Meningitis | | |
|---|---|---|---|---|---|---|
| | Risk Profile | | | Risk Profile | | |
| | Low | Moderate | High | Low | Moderate | High |
| 18-49 | 8.9 | 7.9 | 10.2 | 8.9 | 7.9 | 10.2 |
| 50-59 | 13.5 | 12.0 | 15.6 | 13.5 | 12.0 | 15.6 |
| 60-64 | 12.9 | 11.4 | 14.8 | 12.9 | 11.4 | 14.8 |
| 65-74 | 14.5 | 12.8 | 16.7 | 14.5 | 12.8 | 16.7 |
| 75-99 | 25.6 | 22.7 | 29.4 | 25.6 | 22.7 | 29.4 |

**All-Cause CAP.** Inpatient age- and risk-specific CFRs were based on mortality data from Deb et al. [7], age-specific risk profile from Pelton et al.[3], and risk-specific disease risks for IPD from Deb et al. [7]. In the outpatient setting, the estimates employed in a published Robert Koch Institut analysis were used [10].

**Table 7. Case-fatality risks for inpatient all-cause CAP (per 100)**

| Age Group (years) | Inpatient Care | | | Outpatient Care Only | | |
|---|---|---|---|---|---|---|
| | Risk Profile | | | Risk Profile | | |
| | Low | Moderate | High | Low | Moderate | High |
| 18-49 | 3.9 | 5.9 | 7.5 | 0.0 | 0.0 | 0.0 |
| 50-59 | 8.9 | 13.4 | 17.1 | 0.4 | 0.4 | 0.4 |
| 60-64 | 10.8 | 16.2 | 20.7 | 0.4 | 0.4 | 0.4 |
| 65-74 | 12.2 | 18.3 | 23.5 | 0.4 | 0.4 | 0.4 |
| 75-99 | 13.7 | 20.6 | 26.3 | 0.4 | 0.4 | 0.4 |

#### 2.5 General Population Mortality

For persons projected to be free of IPD and all-cause CAP as well as those who survived IPD and/or inpatient all-cause CAP in a given year, the risk of death was calculated based on age- and risk-specific mortality rates from the general population [11]. Specifically, age-specific general population mortality rates for 2020 (from Destatis) were first allocated across risk groups based on corresponding population weights and relative risks of mortality (assumed to be 1.5 and 2.0 for moderate- and high-risk, respectively, vs. low-risk) [3].

**Table 8. General population mortality rates adjusted for death due to IPD and inpatient all-cause CAP (per 100)**

| Age Group (years) | Risk Profile | | |
|---|---|---|---|
| | Low | Moderate | High |
| 18-49 | 0.1 | 0.1 | 0.1 |
| 50-59 | 0.3 | 0.5 | 0.7 |
| 60-64 | 0.7 | 1.0 | 1.3 |
| 65-74 | 1.2 | 1.8 | 2.4 |
| 75-99 | 4.9 | 7.4 | 9.9 |

#### 2.6 Serotype Distributions

**IPD.**

The proportion of IPD and CAP due to vaccine serotypes is presented in Table 9 and Table 10 respectively. The values for year 1 of the modeling horizon were from unpublished National Reference Laboratory data provided to Pfizer [Pfizer GmbH, data on file]. These were assumed to be stable until the year following the introduction of new PCVs in the pediatric population, after which IPD and CAP attributable to newly-introduced serotypes were assumed to decline. Lacking German data, reductions due to indirect effects from pediatric PCV15/PCV20 vaccination were extrapolated from the reductions in IPD caused by PCV13 non-PCV7 serotypes (excluding serotype 3) observed among adults following the introduction of pediatric PCV13 in the United States (Pfizer Inc, data on file). These reductions are presented in Table 11. These reductions were applied only to the incremental serotypes (i.e., PCV15-non-13 after pediatric introduction of PCV15, and PCV20-non-15 following pediatric introduction of PCV20). We assumed indirect effects of PCV13 were fully realized by year 1 of the model.

**Table 9. Percentage of IPD due to vaccine serotypes, by age and year of modeling horizon**

| Age/Year of Modeling Horizon | PCV13 | PCV15 | PCV20 | PPSV23 | ST 3 | ST 6A/6C | Total Relative Reduction in Disease |
|---|---|---|---|---|---|---|---|
| **18-49** | | | | | | | |
| Year 1 | 27.8% | 34.2% | 68.7% | 78.1% | 12.0% | 1.6% | 0.0% |
| Year 2 | 27.8% | 34.2% | 68.7% | 78.1% | 12.0% | 1.6% | 0.0% |
| Year 3 | 28.4% | 32.7% | 68.0% | 77.6% | 12.3% | 1.6% | 2.2% |
| Year 4 | 33.2% | 35.8% | 62.6% | 73.9% | 14.3% | 1.9% | 16.2% |
| Year 5 | 38.3% | 40.7% | 56.9% | 69.9% | 16.5% | 2.2% | 27.3% |
| Year 6 | 39.9% | 41.6% | 55.1% | 68.6% | 17.2% | 2.3% | 30.3% |
| Year 7 | 41.8% | 43.2% | 52.9% | 67.0% | 18.1% | 2.4% | 33.6% |
| Year 8 | 43.0% | 44.2% | 51.6% | 66.1% | 18.6% | 2.5% | 35.4% |
| Year 9 | 43.7% | 44.6% | 50.8% | 65.6% | 18.9% | 2.5% | 36.4% |
| Year 10 | 44.3% | 45.3% | 50.1% | 65.1% | 19.1% | 2.6% | 37.3% |
| **50-59** | | | | | | | |
| Year 1 | 29.3% | 37.3% | 68.5% | 78.2% | 17.8% | 1.5% | 0.0% |
| Year 2 | 29.3% | 37.3% | 68.5% | 78.2% | 17.8% | 1.5% | 0.0% |
| Year 3 | 29.7% | 36.4% | 68.0% | 77.9% | 18.1% | 1.5% | 1.4% |
| Year 4 | 32.7% | 36.3% | 64.8% | 75.7% | 19.9% | 1.7% | 10.4% |
| Year 5 | 38.6% | 42.1% | 58.5% | 71.3% | 23.4% | 2.0% | 24.1% |
| Year 6 | 40.3% | 42.6% | 56.7% | 70.0% | 24.5% | 2.1% | 27.3% |
| Year 7 | 42.8% | 44.6% | 54.0% | 68.2% | 26.0% | 2.2% | 31.5% |
| Year 8 | 43.8% | 45.5% | 52.9% | 67.4% | 26.6% | 2.2% | 33.1% |
| Year 9 | 44.1% | 45.9% | 52.6% | 67.2% | 26.8% | 2.3% | 33.6% |
| Year 10 | 44.1% | 45.6% | 52.6% | 67.2% | 26.8% | 2.3% | 33.6% |
| **60-64** | | | | | | | |
| Year 1 | 30.3% | 38.9% | 67.0% | 75.3% | 20.8% | 1.8% | 0.0% |
| Year 2 | 30.3% | 38.9% | 67.0% | 75.3% | 20.8% | 1.8% | 0.0% |
| Year 3 | 30.8% | 37.9% | 66.5% | 74.9% | 21.1% | 1.8% | 1.5% |
| Year 4 | 33.7% | 37.6% | 63.3% | 72.5% | 23.2% | 2.0% | 10.2% |
| Year 5 | 39.2% | 42.8% | 57.4% | 68.1% | 26.9% | 2.3% | 22.6% |
| Year 6 | 40.8% | 43.2% | 55.6% | 66.8% | 28.0% | 2.4% | 25.7% |
| Year 7 | 43.0% | 44.9% | 53.2% | 65.0% | 29.5% | 2.6% | 29.5% |
| Year 8 | 43.9% | 45.7% | 52.2% | 64.2% | 30.1% | 2.6% | 31.0% |
| Year 9 | 44.2% | 46.1% | 51.9% | 64.0% | 30.3% | 2.6% | 31.5% |
| Year 10 | 44.2% | 45.7% | 51.8% | 63.9% | 30.4% | 2.6% | 31.5% |
| **65-74** | | | | | | | |
| Year 1 | 30.3% | 38.9% | 67.0% | 75.3% | 20.8% | 2.0% | 0.0% |
| Year 2 | 30.3% | 38.9% | 67.0% | 75.3% | 20.8% | 2.0% | 0.0% |
| Year 3 | 31.1% | 37.2% | 66.1% | 74.6% | 21.4% | 2.1% | 2.7% |
| Year 4 | 35.1% | 39.4% | 61.7% | 71.4% | 24.1% | 2.3% | 13.7% |
| Year 5 | 39.1% | 41.8% | 57.4% | 68.1% | 26.9% | 2.6% | 22.5% |
| Year 6 | 42.5% | 44.2% | 53.7% | 65.3% | 29.2% | 2.8% | 28.7% |
| Year 7 | 44.4% | 45.8% | 51.6% | 63.8% | 30.5% | 2.9% | 31.8% |
| Year 8 | 45.0% | 46.4% | 51.0% | 63.3% | 30.9% | 3.0% | 32.6% |
| Year 9 | 45.0% | 46.3% | 51.0% | 63.3% | 30.9% | 3.0% | 32.7% |
| Year 10 | 45.4% | 46.3% | 50.5% | 63.0% | 31.2% | 3.0% | 33.3% |
| **75-99** | | | | | | | |
| Year 1 | 30.9% | 40.4% | 60.1% | 70.7% | 23.1% | 3.1% | 0.0% |
| Year 2 | 30.9% | 40.4% | 60.1% | 70.7% | 23.1% | 3.1% | 0.0% |
| Year 3 | 31.9% | 38.6% | 58.9% | 69.8% | 23.8% | 3.2% | 3.0% |
| Year 4 | 35.0% | 39.6% | 54.9% | 66.8% | 26.1% | 3.5% | 11.6% |
| Year 5 | 37.9% | 40.7% | 51.1% | 64.1% | 28.3% | 3.8% | 18.4% |
| Year 6 | 40.2% | 41.9% | 48.1% | 61.9% | 30.0% | 4.0% | 23.1% |
| Year 7 | 41.4% | 42.8% | 46.5% | 60.7% | 31.0% | 4.2% | 25.4% |
| Year 8 | 41.7% | 43.2% | 46.1% | 60.4% | 31.2% | 4.2% | 26.0% |
| Year 9 | 41.8% | 43.1% | 46.1% | 60.4% | 31.2% | 4.2% | 26.0% |
| Year 10 | 42.1% | 43.0% | 45.7% | 60.1% | 31.5% | 4.2% | 26.6% |

**All-cause CAP.**

**Table 10. Percentage of all-cause CAP due to vaccine serotypes, by age and year of modeling horizon**

| Age/Year of Modeling Horizon | PCV13 | PCV15 | PCV20 | PPSV23 | ST 3 | ST 6A/6C | | Total Relative Reduction in Disease |
|---|---|---|---|---|---|---|---|---|
| **18-49** | | | | | | | | |
| Year 1 | 10.1% | 11.9% | 14.7% | 14.7% | 1.8% | 1.2% | | 0.0% |
| Year 2 | 10.1% | 11.9% | 14.7% | 14.7% | 1.8% | 1.2% | | 0.0% |
| Year 3 | 10.2% | 11.3% | 14.2% | 14.2% | 1.8% | 1.2% | | 0.6% |
| Year 4 | 10.3% | 11.0% | 12.8% | 12.8% | 1.8% | 1.2% | | 2.2% |
| Year 5 | 10.4% | 10.9% | 11.9% | 11.9% | 1.9% | 1.2% | | 3.1% |
| Year 6 | 10.5% | 10.8% | 11.6% | 11.6% | 1.9% | 1.2% | | 3.5% |
| Year 7 | 10.5% | 10.8% | 11.3% | 11.3% | 1.9% | 1.2% | | 3.8% |
| Year 8 | 10.5% | 10.7% | 11.1% | 11.1% | 1.9% | 1.3% | | 4.0% |
| Year 9 | 10.5% | 10.7% | 11.0% | 11.0% | 1.9% | 1.3% | | 4.1% |
| Year 10 | 10.5% | 10.7% | 11.0% | 11.0% | 1.9% | 1.3% | | 4.2% |
| **50-59** | | | | | | | | |
| Year 1 | 10.1% | 11.9% | 14.7% | 14.7% | 1.8% | 1.2% | | 0.0% |
| Year 2 | 10.1% | 11.9% | 14.7% | 14.7% | 1.8% | 1.2% | | 0.0% |
| Year 3 | 10.1% | 11.6% | 14.4% | 14.4% | 1.8% | 1.2% | | 0.3% |
| Year 4 | 10.3% | 11.0% | 13.3% | 13.3% | 1.8% | 1.2% | | 1.6% |
| Year 5 | 10.4% | 11.0% | 12.2% | 12.2% | 1.9% | 1.2% | | 2.9% |
| Year 6 | 10.4% | 10.8% | 11.8% | 11.8% | 1.9% | 1.2% | | 3.3% |
| Year 7 | 10.5% | 10.8% | 11.4% | 11.4% | 1.9% | 1.2% | | 3.7% |
| Year 8 | 10.5% | 10.8% | 11.2% | 11.2% | 1.9% | 1.2% | | 3.9% |
| Year 9 | 10.5% | 10.8% | 11.2% | 11.2% | 1.9% | 1.2% | | 3.9% |
| Year 10 | 10.5% | 10.7% | 11.2% | 11.2% | 1.9% | 1.2% | | 4.0% |
| **60-64** | | | | | | | | |
| Year 1 | 7.2% | 8.0% | 12.6% | 15.0% | 3.8% | 0.0% | | 0.0% |
| Year 2 | 7.2% | 8.0% | 12.6% | 15.0% | 3.8% | 0.0% | | 0.0% |
| Year 3 | 7.2% | 7.9% | 12.5% | 14.9% | 3.8% | 0.0% | | 0.1% |
| Year 4 | 7.3% | 7.6% | 11.4% | 13.9% | 3.9% | 0.0% | | 1.3% |
| Year 5 | 7.4% | 7.7% | 9.6% | 12.1% | 3.9% | 0.0% | | 3.3% |
| Year 6 | 7.5% | 7.7% | 9.2% | 11.7% | 3.9% | 0.0% | | 3.7% |
| Year 7 | 7.5% | 7.7% | 8.7% | 11.2% | 4.0% | 0.0% | | 4.3% |
| Year 8 | 7.5% | 7.7% | 8.4% | 10.9% | 4.0% | 0.0% | | 4.6% |
| Year 9 | 7.5% | 7.7% | 8.4% | 10.9% | 4.0% | 0.0% | | 4.6% |
| Year 10 | 7.5% | 7.6% | 8.4% | 10.9% | 4.0% | 0.0% | | 4.6% |
| **65-74** | | | | | | | | |
| Year 1 | 7.2% | 8.0% | 12.6% | 15.0% | 3.8% | 0.0% | | 0.0% |
| Year 2 | 7.2% | 8.0% | 12.6% | 15.0% | 3.8% | 0.0% | | 0.0% |
| Year 3 | 7.2% | 7.8% | 12.4% | 14.8% | 3.8% | 0.0% | | 0.3% |
| Year 4 | 7.3% | 7.7% | 10.9% | 13.4% | 3.9% | 0.0% | | 1.9% |
| Year 5 | 7.4% | 7.6% | 9.7% | 12.2% | 3.9% | 0.0% | | 3.2% |
| Year 6 | 7.5% | 7.6% | 8.8% | 11.3% | 4.0% | 0.0% | | 4.2% |
| Year 7 | 7.6% | 7.6% | 8.3% | 10.8% | 4.0% | 0.0% | | 4.7% |
| Year 8 | 7.6% | 7.7% | 8.2% | 10.7% | 4.0% | 0.0% | | 4.8% |
| Year 9 | 7.6% | 7.6% | 8.2% | 10.7% | 4.0% | 0.0% | | 4.8% |
| Year 10 | 7.6% | 7.6% | 8.1% | 10.6% | 4.0% | 0.0% | | 4.9% |
| **75-99** | | | | | | | | |
| Year 1 | 7.2% | 8.0% | 12.6% | 15.0% | 3.8% | 0.0% | | 0.0% |
| Year 2 | 7.2% | 8.0% | 12.6% | 15.0% | 3.8% | 0.0% | | 0.0% |
| Year 3 | 7.2% | 7.8% | 12.4% | 14.8% | 3.8% | 0.0% | | 0.3% |
| Year 4 | 7.3% | 7.7% | 10.9% | 13.4% | 3.9% | 0.0% | | 1.9% |
| Year 5 | 7.4% | 7.6% | 9.7% | 12.2% | 3.9% | 0.0% | | 3.2% |
| Year 6 | 7.5% | 7.6% | 8.8% | 11.3% | 4.0% | 0.0% | | 4.2% |
| Year 7 | 7.6% | 7.6% | 8.3% | 10.8% | 4.0% | 0.0% | | 4.7% |
| Year 8 | 7.6% | 7.7% | 8.2% | 10.7% | 4.0% | 0.0% | | 4.8% |
| Year 9 | 7.6% | 7.6% | 8.2% | 10.7% | 4.0% | 0.0% | | 4.8% |
| Year 10 | 7.6% | 7.6% | 8.1% | 10.6% | 4.0% | 0.0% | | 4.9% |

**Table 11. Relative reductions in vaccine-type IPD or CAP due to indirect effects of pediatric PCV immunization with PCV15 or PCV20, by age and year of use**

| Years since pediatric introduction | 18-49 | 50-59 | 60-64 | 65-74 | 75 and older |
|---|---|---|---|---|---|
| 1 | 0.0% | 0.0% | 0.0% | 0.0% | 0.0% |
| 2 | 34.8% | 17.9% | 17.9% | 31.5% | 31.5% |
| 3 | 65.7% | 59.9% | 59.9% | 57.0% | 57.0% |
| 4 | 72.7% | 67.1% | 67.1% | 76.0% | 76.0% |
| 5 | 81.3% | 79.3% | 79.3% | 85.9% | 85.9% |
| 6 | 86.1% | 84.1% | 84.1% | 89.0% | 89.0% |
| 7 | 88.6% | 85.9% | 85.9% | 88.8% | 88.8% |
| 8 | 91.4% | 85.1% | 85.1% | 90.0% | 90.0% |
| 9 | 90.1% | 88.2% | 88.2% | 93.1% | 93.1% |
| 10 | No further reduction – serotype distribution stable until end of model | | | | |

#### 2.7 Vaccine Effectiveness

For each vaccine and each clinical manifestation of pneumococcal disease, vaccine effectiveness (VE) against vaccine-type (VT) disease was estimated by age in one-year increments for all adults aged 18-99 years, for the first year following receipt (i.e., first year of the modeling horizon) and all subsequent years, respectively.

##### 2.7.1 VE-PCV vs. VT-IPD

Effectiveness of PCV against vaccine-type IPD (VT-IPD) for low-risk and moderate-risk persons in year 1 of the modeling horizon ("initial VE") was based on data for adults aged ≥65 years in the CAPiTA per-protocol population, and from post-hoc analyses of CAPiTA data extrapolated for adults aged <65 years [12,13]. For low-/moderate-risk persons aged ≥65 years, initial VE was assumed to be 75.0% based on overall findings from the per-protocol population in CAPiTA [12]. For low-/moderate-risk persons aged 50-64 years, initial VE by single year of age was derived using age-specific relative changes in VE against VT-IPD (vs. age 65 years) from Mangen et al. [13]. For persons aged 18-49 years, initial VE was assumed to be the same as that for persons aged 50 years [13].

Initial VE against VT-IPD for high-risk persons aged 18-99 years was assumed to be equal to 80% of corresponding values for low-/moderate-risk. This assumption was based on the study by Klugman et al. in which PCV9 vaccine efficacy against VT-IPD among HIV+ children was 65%, versus 83% among HIV- children (relative efficacy = 78%).[14] The finding of high vaccine efficacy among immunocompromised persons was also supported by findings from the study by French et al. in which efficacy of PCV7 (2 doses) against VT-IPD was 74% among HIV+ adults [15].

Initial VE was assumed to persist for the first 5 years of the modeling horizon for all ages and risk groups, consistent with CAPiTA, in which no waning of vaccine efficacy was observed during the 5-year follow-up period [12,16]. Beyond year 5 of the modeling horizon, VE was assumed to wane across all age and risk groups—based on assumptions employed by CAPiTA investigators (i.e., Mangen et al.)—as follows: 5% annual decline during years 6-10, 10% annual decline during years 11-15, and no efficacy from year 16 through the end of the modeling horizon [13]. VE was assumed to be the same across vaccine-specific serotypes.

**Table 12. Effectiveness of PCV against vaccine-type IPD**

| Age Group (years)/Risk | Year 1 | Year 5 | Year 10 | Year 15 | Year 16+ |
|---|---|---|---|---|---|
| 18-49 | | | | | |
| Low | 81.5% | 81.5% | 63.1% | 37.2% | 0.0% |
| Moderate | 81.5% | 81.5% | 63.1% | 37.2% | 0.0% |
| High | 65.2% | 65.2% | 50.5% | 29.8% | 0.0% |
| 50-59 | | | | | |
| Low | 80.1% | 80.1% | 62.0% | 36.6% | 0.0% |
| Moderate | 80.0% | 80.0% | 61.9% | 36.6% | 0.0% |
| High | 63.9% | 63.9% | 49.5% | 29.2% | 0.0% |
| 60-64 | | | | | |
| Low | 76.9% | 76.9% | 59.5% | 35.1% | 0.0% |
| Moderate | 76.9% | 76.9% | 59.5% | 35.1% | 0.0% |
| High | 61.5% | 61.5% | 47.6% | 28.1% | 0.0% |
| 65-74 | | | | | |
| Low | 75.0% | 75.0% | 58.0% | 34.3% | 0.0% |
| Moderate | 75.0% | 75.0% | 58.0% | 34.3% | 0.0% |
| High | 60.0% | 60.0% | 46.4% | 27.4% | 0.0% |
| 75-99 | | | | | |
| Low | 75.0% | 75.0% | 58.0% | 34.3% | 0.0% |
| Moderate | 75.0% | 75.0% | 58.0% | 34.3% | 0.0% |
| High | 60.0% | 60.0% | 46.4% | 27.4% | 0.0% |

##### 2.7.2 VE-PPSV23 vs. VT-IPD

Effectiveness of PPSV23 against VT-IPD for low-risk, moderate-risk, and high-risk persons aged 18 years and older in year 1 of the modeling horizon was estimated using data from the published literature[17,18]. Initial VE was derived for all ages by fitting a logarithmic curve to values for persons aged 65-74, 75-84, and 85-99 years, respectively, from Djennad et al., and then allocating estimated age-specific values across risk groups using relative risks from Djennad et al. and corresponding population weights[17,18].

Beyond year 1 of the modeling horizon, VE was assumed to wane across all age and risk groups as follows: linear decline to 76.2% of initial VE by year 5, and linear decline to no efficacy by year 10 and through the end of the modeling horizon [19]. VE was assumed to be the same across vaccine-specific serotypes.

**Table 13. Effectiveness of PPSV23 against VT-IPD**

| Age Group (years)/Risk | Year 1 | Year 5 | Year 10 | Year 15 | Year 16+ |
|---|---|---|---|---|---|
| 18-49 | | | | | |
| Low | 59.1% | 45.1% | 0.0% | 0.0% | 0.0% |
| Moderate | 32.8% | 25.0% | 0.0% | 0.0% | 0.0% |
| High | 17.1% | 13.0% | 0.0% | 0.0% | 0.0% |
| 50-59 | | | | | |
| Low | 58.6% | 44.6% | 0.0% | 0.0% | 0.0% |
| Moderate | 32.5% | 24.8% | 0.0% | 0.0% | 0.0% |
| High | 16.9% | 12.9% | 0.0% | 0.0% | 0.0% |
| 60-64 | | | | | |
| Low | 57.6% | 43.9% | 0.0% | 0.0% | 0.0% |
| Moderate | 32.0% | 24.4% | 0.0% | 0.0% | 0.0% |
| High | 16.6% | 12.7% | 0.0% | 0.0% | 0.0% |
| 65-74 | | | | | |
| Low | 55.8% | 42.5% | 0.0% | 0.0% | 0.0% |
| Moderate | 30.9% | 23.6% | 0.0% | 0.0% | 0.0% |
| High | 16.0% | 12.2% | 0.0% | 0.0% | 0.0% |
| 75-99 | | | | | |
| Low | 48.2% | 36.7% | 0.0% | 0.0% | 0.0% |
| Moderate | 25.7% | 19.6% | 0.0% | 0.0% | 0.0% |
| High | 13.1% | 10.0% | 0.0% | 0.0% | 0.0% |

##### 2.7.3 VE-PCV vs. VT-CAP

Effectiveness of PCV against vaccine-type CAP (VT-CAP) for low-risk and moderate-risk persons in year 1 of the modeling horizon ("initial VE") was based on data for adults aged ≥65 years in the CAPiTA per-protocol population, and from post-hoc analyses of CAPiTA data extrapolated for adults <65 years[12,13]. For low-/moderate-risk persons aged ≥65 years, initial VE was assumed to be 45.0% based on overall findings from the per-protocol population in CAPiTA [12]. For low-/moderate-risk persons aged 50-64 years, initial VE by single year of age was derived using age-specific relative changes in VE against VT-CAP (vs. age 65 years) from Mangen et al. [13]. For persons aged 18-49 years, initial VE was assumed to be the same as that for persons aged 50 years.

Initial VE against VT-CAP for high-risk persons aged 18-99 years was assumed to be equal to 80% of corresponding values for low-/moderate-risk persons based on the same adjustment factor used for VT-IPD in the high-risk. Data on the real-world effectiveness of PCV13 among adults also showed little change in VT-CAP effectiveness after adjustment for patient risk profile, suggesting that employing a downward adjustment factor for VE among high-risk adults (vs. low-/moderate-risk adults) is likely conservative [20].

Initial VE was assumed to persist for the first 5 years of the modeling horizon for all ages and risk groups, consistent with CAPiTA, in which no waning of vaccine efficacy was observed during the 5-year follow-up period [12,16]. Beyond year 5 of the modeling horizon, VE was assumed to wane across all age and risk groups—based on assumptions employed by CAPiTA investigators (i.e., Mangen et al.)—as follows: 5% annual decline during years 6-10, 10% annual decline during years 11-15, and no efficacy from year 16 through the end of the modeling horizon [13]. VE was assumed to be the same across vaccine-specific serotypes.

**Table 14. Effectiveness of PCV against VT-CAP**

| Age Group (years)/Risk | Year 1 | Year 5 | Year 10 | Year 15 | Year 16+ |
|---|---|---|---|---|---|
| 18-49 | | | | | |
| Low | 55.6% | 55.6% | 43.0% | 25.4% | 0.0% |
| Moderate | 55.6% | 55.6% | 43.0% | 25.4% | 0.0% |
| High | 44.5% | 44.5% | 34.4% | 20.3% | 0.0% |
| 50-59 | | | | | |
| Low | 53.0% | 53.0% | 41.0% | 24.2% | 0.0% |
| Moderate | 52.8% | 52.8% | 40.8% | 24.1% | 0.0% |
| High | 42.1% | 42.1% | 32.6% | 19.2% | 0.0% |
| 60-64 | | | | | |
| Low | 47.6% | 47.6% | 36.8% | 21.8% | 0.0% |
| Moderate | 47.5% | 47.5% | 36.8% | 21.7% | 0.0% |
| High | 38.0% | 38.0% | 29.4% | 17.3% | 0.0% |
| 65-74 | | | | | |
| Low | 45.0% | 45.0% | 34.8% | 20.6% | 0.0% |
| Moderate | 45.0% | 45.0% | 34.8% | 20.6% | 0.0% |
| High | 36.0% | 36.0% | 27.9% | 16.4% | 0.0% |
| 75-99 | | | | | |
| Low | 45.0% | 45.0% | 34.8% | 20.6% | 0.0% |
| Moderate | 45.0% | 45.0% | 34.8% | 20.6% | 0.0% |
| High | 36.0% | 36.0% | 27.9% | 16.4% | 0.0% |

###### 2.7.4 VE-PPSV23 vs. VT-CAP

Effectiveness of PPSV23 against VT-CAP was assumed to be zero based on various published sources, and consistent with base-case assumptions employed in a number of economic studies[21-37].

##### 2.8 Health State Utility Values

General population health state utilities were the same as those employed in a previously published cost-effectiveness analysis of pneumococcal vaccination among German adults [38].

**Table 15. Annual general population health-state utility values**

| Age Group (years) | Risk Profile | | |
|---|---|---|---|
| | Low | Moderate | High |
| 18-49 | 0.948 | 0.812 | 0.812 |
| 50-59 | 0.884 | 0.729 | 0.729 |
| 60-64 | 0.849 | 0.705 | 0.705 |
| 65-74 | 0.834 | 0.662 | 0.662 |
| 75-99 | 0.814 | 0.568 | 0.568 |

##### 2.9 Utility Decrements

Disutilities due to IPD and inpatient all-cause CAP (0.130) were based on the utility difference between patients with and without suspected pneumonia at one-year post-discharge (from pneumonia hospitalization) reported by Mangen et al. [39]. Utility values reported by Mangen et al. were derived using an area under the curve approach [39]. Disutilities due to all-cause CAP requiring outpatient care only (0.004) were based on data from a study by Melegaro et al. [40]. All disutilities were assumed to be the same irrespective of age and risk.

##### 2.10 Vaccine Costs

The prices of PPSV23 (33.88 €), PCV13 (72.50 €), PCV15 (76.74 €), and PCV20 (76.74 €) were based on average retail pharmacy price per dose [41]. The cost of vaccine administration was assumed to be 8.19 € .[38] All costs are reported in 2022 Euro.

##### 2.11 Medical Care Costs

Age-specific costs of IPD hospitalization (bacteremia and meningitis, respectively) were from a previously published cost-effectiveness analysis of pneumococcal vaccination in Germany [38]. Age- and risk-specific costs of all-cause CAP requiring hospitalization and outpatient care only, respectively, were derived from Pfizer data on file and included all-cause healthcare expenditures within 30 days of the start of the CAP episode (i.e., date of hospital admission or date of outpatient encounter for CAP) [42].

**Table 16. Medical care costs for IPD (per case)**

| Age Group (years) | Bacteremia | | | Meningitis | | |
|---|---|---|---|---|---|---|
| | Risk Profile | | | Risk Profile | | |
| | Low | Moderate | High | Low | Moderate | High |
| 18-49 | 9,397 € | 9,397 € | 9,397 € | 4,732 € | 4,732 € | 4,732 € |
| 50-59 | 10,097 € | 10,097 € | 10,097 € | 5,129 € | 5,129 € | 5,129 € |
| 60-64 | 9,928 € | 9,928 € | 9,928 € | 5,260 € | 5,260 € | 5,260 € |
| 65-74 | 7,773 € | 7,773 € | 7,773 € | 5,326 € | 5,326 € | 5,326 € |
| 75-99 | 5,326 € | 5,326 € | 5,326 € | 5,348 € | 5,348 € | 5,348 € |

**Table 17. Medical care costs for all-cause CAP (per case)**

| Age Group (years) | Inpatient Care | | | Outpatient Care Only | | |
|---|---|---|---|---|---|---|
| | Risk Profile | | | Risk Profile | | |
| | Low | Moderate | High | Low | Moderate | High |
| 18-49 | 5,261 € | 7,375 € | 9,900 € | 361 € | 563 € | 1,214 € |
| 50-59 | 8,213 € | 8,604 € | 8,709 € | 542 € | 706 € | 1,233 € |
| 60-64 | 7,186 € | 9,584 € | 9,859 € | 535 € | 745 € | 1,931 € |
| 65-74 | 6,734 € | 9,529 € | 8,194 € | 763 € | 901 € | 1,949 € |
| 75-99 | 6,108 € | 6,753 € | 6,682 € | 1,003 € | 1,227 € | 1,870 € |

##### 2.12 Discounting

Costs and life years were discounted at a 3.0% annual rate.

##### 2.13 Analytic Perspective

Analyses were conducted from the payer perspective (i.e., excluding patient paid amounts). All costs were expressed in 2022 Euros (€).

##### 2.14 Modeling Horizon

Clinical outcomes and economic costs were evaluated for the model populations on an annual basis, from model entry through end of life.

#### 3 Estimation of Model Inputs for Scenario Analysis

##### 3.1 PCV20 vs. PCV15→PPSV23

###### 3.1.1 Population Size

The model population included was the same as in base case analyses (Section 2.1).

###### 3.1.2 Vaccine Coverage

For persons receiving PCV20 alone, vaccine coverage was estimated using the same sources as for base cases analyses (Section 2.2); coverage levels are detailed in Table 18.

**Table 18. Vaccine coverage with PCV20**

| Age Group (years) | Risk Profile | | |
|---|---|---|---|
| | Low | Moderate | High |
| 18-49 | 0.0% | 7.1% | 7.1% |
| 50-59 | 0.0% | 12.5% | 12.5% |
| 60-64 | 13.4% | 31.8% | 31.8% |
| 65-74 | 34.2% | 35.1% | 35.1% |
| 75-99 | 41.9% | 35.2% | 35.2% |

For persons receiving PCV15→PPSV23, vaccine coverage was estimated using the same sources as for base case analyses; coverage levels are detailed in Table 19. All persons who received PCV15 at model entry and who were alive at the start of year 2 were assumed to receive PPSV23.

**Table 19. Vaccine coverage with PCV15→PPSV23**

| Age Group (years) | Risk Profile | | |
|---|---|---|---|
| | Low | Moderate | High |
| 18-49 | 0.0% | 7.1% | 7.1% |
| 50-59 | 0.0% | 12.5% | 12.5% |
| 60-64 | 13.4% | 31.8% | 31.8% |
| 65-74 | 34.2% | 35.1% | 35.1% |
| 75-99 | 41.9% | 35.2% | 35.2% |

### 4 Estimation of Model Inputs for Sensitivity Analyses

The sensitivity of the cost-effectiveness of PCV20 due to changes in key model parameter values and assumptions was explored. The following were increased and decreased by 25% (relative to the base case):

- Incidence of bacteremia
- Incidence of meningitis
- Incidence of inpatient all cause CAP
- Incidence of outpatient all cause CAP
- Case fatality risk of bacteremia
- Case fatality risk of meningitis
- Case fatality risk of inpatient all cause CAP
- Case fatality risk of outpatient all cause CAP
- Cost of bacteremia
- Cost of meningitis
- Cost of inpatient all cause CAP
- Cost of outpatient all cause CAP
- All cost
- PPSV23 VE-IPD
- PCV13/PCV20 VE-IPD
- PCV13/PCV20 VE-CAP
- General population utility
- Disutility of bacteremia
- Disutility of meningitis
- Disutility of inpatient all cause CAP
- Disutility of outpatient all cause CAP
- Herd effects on IPD
- Herd effects on CAP

Further sensitivity analyses were conducted by exploring the following

- 10% revaccination with PPSV23 (SC only)

Assuming the proportion of persons receiving SC who were revaccinated with PPSV23 was reduced to 10% instead of 100% (base case).

- No herd effects
  Assuming herd effects from PCV20 and PCV15 use among infants and children not occurring. Thus, overall levels of disease and the proportion of disease due to vaccine serotypes remained constant throughout the modelling horizon.
- Full replacement of serotypes
  Assuming that VT-serotypes were fully replaced.
- VE-PPSV23 vs. VT-CAP from Suzuki et al. [19]
- VE-PPSV23 vs. VT-CAP from Lawrence et al. [43]
- VE-PPSV23 vs. VT-CAP from Lawrence et al. [43] with 10-year waning

##### 4.1 VE-PPSV23 vs. VT-CAP Based on Suzuki et al.

Effectiveness of PPSV23 against VT-CAP was based principally on Suzuki et al. and employed other published literature, as needed[17,19]. Initial VE for persons aged 75-84 years was based on adjusted vaccine effectiveness (37.7%, during 1-month to 2-year follow-up period) reported by Suzuki et al., which was anchored to low-risk persons aged 79 years [19]. (Age 79 years was chosen as the anchor point because it is the midpoint of the 75-84-year-old age group, which was the largest age group in the Suzuki et al. study.) Initial VE for low-risk persons by single year of age was derived using age-specific relative changes in VE against VT-IPD (vs. persons aged 79 years) based on Djennad et al.19 Initial VE for moderate- and high-risk persons by single year of age was derived using age- and risk-specific relative changes in VE against VT-IPD (vs. low-risk) based on Djennad et al. [17]. Although the point estimate reported by Suzuki et al. for VE ≥5 years after receipt of vaccination was 26%, the confidence interval ranged from -56% to 65%, indicating a high degree of uncertainty.[19] Thus, lacking more robust data, VE was assumed to wane linearly—across all age and risk groups—to 0% within 5 years of vaccination.[19]

**Table 20. Effectiveness of PPSV23 against VT-CAP based on Suzuki et al.**

| Age Group (years)/Risk | Year 1 | Year 5 | Year 10 | Year 15 | Year 16+ |
|---|---|---|---|---|---|
| 18-49 | | | | | |
| Low | 43.8% | 0.0% | 0.0% | 0.0% | 0.0% |
| Moderate | 24.3% | 0.0% | 0.0% | 0.0% | 0.0% |
| High | 12.6% | 0.0% | 0.0% | 0.0% | 0.0% |
| 50-59 | | | | | |
| Low | 43.8% | 0.0% | 0.0% | 0.0% | 0.0% |
| Moderate | 24.3% | 0.0% | 0.0% | 0.0% | 0.0% |
| High | 12.7% | 0.0% | 0.0% | 0.0% | 0.0% |
| 60-64 | | | | | |
| Low | 43.8% | 0.0% | 0.0% | 0.0% | 0.0% |
| Moderate | 24.3% | 0.0% | 0.0% | 0.0% | 0.0% |
| High | 12.7% | 0.0% | 0.0% | 0.0% | 0.0% |
| 65-74 | | | | | |
| Low | 43.8% | 0.0% | 0.0% | 0.0% | 0.0% |
| Moderate | 24.3% | 0.0% | 0.0% | 0.0% | 0.0% |
| High | 12.7% | 0.0% | 0.0% | 0.0% | 0.0% |
| 75-99 | | | | | |
| Low | 43.8% | 0.0% | 0.0% | 0.0% | 0.0% |
| Moderate | 24.3% | 0.0% | 0.0% | 0.0% | 0.0% |
| High | 12.7% | 0.0% | 0.0% | 0.0% | 0.0% |

##### 4.2 VE-PPSV23 vs. VT-CAP Based on Lawrence et al.

Effectiveness of PPSV23 against VT-CAP was based principally on Lawrence et al. and employed other published literature, as needed [17-19,43]. Initial VE was derived for all ages by fitting a logarithmic curve to VE for persons aged 18-74 and ≥75 years, respectively, from Lawrence et al., and then allocating estimated age-specific values across risk groups using relative risks from Djennad et al. and corresponding population weights [17,18,43]. Initial VE was assumed to wane linearly—across all age and risk groups—to 0% within 5 years of vaccination, based on Suzuki et al. (described above) [19].

**Table 21. Effectiveness of PPSV23 against VT-CAP based on Lawrence et al.**

| Age Group (years)/Risk | Year 1 | Year 5 | Year 10 | Year 15 | Year 16+ |
|---|---|---|---|---|---|
| 18-49 | | | | | |
| Low | 79.2% | 0.0% | 0.0% | 0.0% | 0.0% |
| Moderate | 48.8% | 0.0% | 0.0% | 0.0% | 0.0% |
| High | 26.1% | 0.0% | 0.0% | 0.0% | 0.0% |
| 50-59 | | | | | |
| Low | 79.2% | 0.0% | 0.0% | 0.0% | 0.0% |
| Moderate | 48.8% | 0.0% | 0.0% | 0.0% | 0.0% |
| High | 26.1% | 0.0% | 0.0% | 0.0% | 0.0% |
| 60-64 | | | | | |
| Low | 79.2% | 0.0% | 0.0% | 0.0% | 0.0% |
| Moderate | 48.8% | 0.0% | 0.0% | 0.0% | 0.0% |
| High | 26.1% | 0.0% | 0.0% | 0.0% | 0.0% |
| 65-74 | | | | | |
| Low | 79.2% | 0.0% | 0.0% | 0.0% | 0.0% |
| Moderate | 48.8% | 0.0% | 0.0% | 0.0% | 0.0% |
| High | 26.1% | 0.0% | 0.0% | 0.0% | 0.0% |
| 75-99 | | | | | |
| Low | 79.2% | 0.0% | 0.0% | 0.0% | 0.0% |
| Moderate | 48.8% | 0.0% | 0.0% | 0.0% | 0.0% |
| High | 26.1% | 0.0% | 0.0% | 0.0% | 0.0% |

##### 4.3 VE-PPSV23 vs. VT-CAP from Lawrence et al. with 10-Year Waning

Initial effectiveness of PPSV23 against VT-CAP was based principally on Lawrence et al. (see Section 4.2). After model year 1, VE was assumed to wane linearly to 0% by model year 10 (based on assumption).

**Table 22. Effectiveness of PPSV23 against VT-CAP based on Lawrence et al. with linear waning over 10 years**

| Age Group (years)/Risk | Year 1 | Year 5 | Year 10 | Year 15 | Year 16+ |
|---|---|---|---|---|---|
| 18-49 | | | | | |
| Low | 79.8% | 44.4% | 0.0% | 0.0% | 0.0% |
| Moderate | 44.5% | 24.7% | 0.0% | 0.0% | 0.0% |
| High | 23.2% | 12.9% | 0.0% | 0.0% | 0.0% |
| 50-59 | | | | | |
| Low | 79.8% | 44.4% | 0.0% | 0.0% | 0.0% |
| Moderate | 44.5% | 24.7% | 0.0% | 0.0% | 0.0% |
| High | 23.2% | 12.9% | 0.0% | 0.0% | 0.0% |
| 60-64 | | | | | |
| Low | 79.8% | 44.4% | 0.0% | 0.0% | 0.0% |
| Moderate | 44.5% | 24.7% | 0.0% | 0.0% | 0.0% |
| High | 23.2% | 12.9% | 0.0% | 0.0% | 0.0% |
| 65-74 | | | | | |
| Low | 79.8% | 44.4% | 0.0% | 0.0% | 0.0% |
| Moderate | 44.5% | 24.7% | 0.0% | 0.0% | 0.0% |
| High | 23.2% | 12.9% | 0.0% | 0.0% | 0.0% |
| 75-99 | | | | | |
| Low | 79.8% | 44.4% | 0.0% | 0.0% | 0.0% |
| Moderate | 44.5% | 24.7% | 0.0% | 0.0% | 0.0% |
| High | 23.2% | 12.9% | 0.0% | 0.0% | 0.0% |

### 5 Estimation of Model Inputs for Scenario Analyses

The sensitivity of the cost-effectiveness of PCV20 due to changes in in a combination of parameter values and assumptions was explored. The following scenarios were tested:

- Comparison of PCV20 and PCV15→PPSV23 (without PPSV23 revaccination)
- Societal perspective
- Vaccine effectiveness parameters (including waning) were changed using sources from independent bodies such as CDC
- Case fatality risks were changed using several different sources
- Lowest PCV20 efficacy assumptions and highest cost assumptions were combined.

#### 5.1 Societal Perspective

The value of work loss due to disease-related morbidity for persons experiencing illness was estimated based on the number of work-loss days due to disease, work-force participation rates, and average daily wage. The average number of work-loss days was estimated based on findings from an ongoing Pfizer study estimating the cost of CAP [Pfizer GmbH, data on file]. The age-specific percentage of persons in the work force and average value of a day of work (i.e., daily wage) were based on published data from Destatis [44,45].

**Table 23. Morbidity-related work-loss days per person in the work force with disease**

| Age Group (years) | IPD | | |
|---|---|---|---|
| | Risk Profile | | |
| | Low | Moderate | High |
| 18-49 | 16 | 18 | 18 |
| 50-59 | 18 | 18 | 18 |
| 60-64 | 17 | 18 | 16 |

| | | | |
|---|---|---|---|
| 65-74 | 0 | 0 | 0 |
| 75-99 | 0 | 0 | 0 |

#### 5.2 Vaccine Effectiveness from US CDC Analysis

A scenario analysis was conducted using the vaccine effectiveness values and waning assumed in the cost-effectiveness model for Japan and the United States CDC cost-effectiveness model presented to the Advisory Committee on Immunization Practices in September of 2021 during their evaluation of PCV20 for adults [46]. These values are presented in the table below. Unlike the base-case effectiveness inputs, these do not vary by age.

**Table 24. CDC vaccine effectiveness and waning assumptions for scenario analysis**

| Parameter | PCV | PPSV23 |
|---|---|---|
| Duration / waning of protection | 15 years:<br>Stable for 5 years<br>Linear decline to 0% over following 10 years | 15 years:<br>Linear decline to 0% over 15 years |
| VE vs VT-IPD (non-ST3) | Low Risk: 75%<br>Moderate Risk: 75%<br>High Risk: 25% | Low Risk: 59.7%<br>Moderate Risk: 59.7%<br>High Risk: 7.9% |
| VE vs ST3 IPD | Low Risk: 26%<br>Moderate Risk: 26%<br>High Risk: 8.7% | Low Risk: 59.7%<br>Moderate Risk: 59.7%<br>High Risk: 7.9% |
| VE vs VT-NBP (non-ST3) | Low Risk: 66.7%<br>Moderate Risk: 40.3%<br>High Risk: 15.0% | Low Risk: 20.0%<br>Moderate Risk: 20.0%<br>High Risk: 6.7% |
| VE vs ST3 NBP | Low & Moderate risk: 15.6%<br>Moderate Risk: 15.6%<br>High Risk: 5.2% | Low Risk: 20.0%<br>Moderate Risk: 20.0%<br>High Risk: 6.7% |

#### 5.3 Case fatality risks

Scenario analyses was conducted using the case fatality risk values from different sources. First, we used alternative German CAP-CFR published by Kolditz and colleagues and IPD-CFR published by van Hoek, which are widely referenced. However, they are less conservative than the base-case values. Second, we used fatality risks that were used in a cost-effectiveness analysis that was conducted for the Robert Koch Institut to evaluate several vaccination alternatives in Germany in 2016. Most of these estimates were more conservative.

**Table 25. Case-fatality risks based on widely published German publications**

| Age group (merged) | IPD (per 100) [18] | | | Inpatient CAP [47] | | | Outpatient CAP [47] | | |
|---|---|---|---|---|---|---|---|---|---|
| | Low | Moderate | High | Low | Moderate | High | Low | Moderate | High |
| 18-49 | 5.4 | 18.2 | 15.4 | 3.3 | 5.0 | 6.4 | 0.1 | 0.1 | 0.1 |
| 50-59 | 5.4 | 18.2 | 15.4 | 9.5 | 14.3 | 18.3 | 0.4 | 0.6 | 0.8 |
| 60-64 | 5.4 | 18.2 | 15.4 | 11.8 | 17.8 | 22.8 | 1.1 | 1.7 | 2.2 |
| 65-74 | 29.1 | 33.0 | 29.9 | 12.3 | 18.5 | 23.7 | 2.0 | 3.0 | 3.8 |
| 75-99 | 29.1 | 33.0 | 29.9 | 17.6 | 26.5 | 33.9 | 6.8 | 10.3 | 13.2 |

**Table 26. Case-fatality risks based on widely published German publications [10]**

| Age group (merged) | IPD (per 100) | | | Inpatient CAP | | | Outpatient CAP | | |
|---|---|---|---|---|---|---|---|---|---|
| | Low | Moderate | High | Low | Moderate | High | Low | Moderate | High |
| 18-49 | 5.4 | 18.2 | 15.4 | 2.1 | 1.8 | 2.4 | - | - | - |
| 50-59 | 5.4 | 18.2 | 15.4 | 6.2 | 5.5 | 7.1 | 0.4 | 0.4 | 0.4 |
| 60-64 | 5.4 | 18.2 | 15.4 | 6.2 | 5.5 | 7.1 | 0.4 | 0.4 | 0.4 |
| 65-74 | 29.1 | 33.0 | 29.9 | 10.2 | 9.1 | 11.8 | 0.4 | 0.4 | 0.4 |
| 75-99 | 29.1 | 33.0 | 29.9 | 16.0 | 14.2 | 18.4 | 0.4 | 0.4 | 0.4 |

##### 5.4 Alternative cost estimates

In one scenario, we evaluated the impact of alternative medical costs by using costs from earlier years (2014) that we used for a cost-effectiveness analysis conducted for the Robert Koch Institute (RKI) which is a government's central scientific institution in Germany. These costs were similar for bacteremia, higher for meningitis, lower for in- and outpatient CAP than the costs used in the base case.

**Table 27: Costs based on cost effectiveness model for the RKI**

| Disease/age groups | Cost per case in Euro |
|---|---|
| IPD all ages | 9,524.91 |
| CAP inpat all ages | 3,527.58 |
| CAP outpat | |
| 60-74 | 44.20 |
| 75+ | 50.16 |

#### 6 Sensitivity analyses Results

The table below show sensitivity results not shown in the manuscript.

**Table 28. Additional Sensitivity Analyses**

| Sensitivity analyses | Difference in Costs (millions) | Difference in LYs | Difference in QALYs | Incremental Cost per LY |
|---|---|---|---|---|
| Base Case | -356.70 € | 74,694 | 49,655 | Dominant |
| base case general population utility (-25%) | -356.70 € | 74,694 | 38,499 | Dominant |
| base case general population utility (+25%) | -356.70 € | 74,694 | 60,680 | Dominant |
| base case bacteremia disutility (-25%) | -356.70 € | 74,694 | 49,594 | Dominant |
| base case bacteremia disutility (+25%) | -356.70 € | 74,694 | 49,716 | Dominant |
| base case meningitis disutility (-25%) | -356.70 € | 74,694 | 49,653 | Dominant |
| base case meningitis disutility (+25%) | -356.70 € | 74,694 | 49,657 | Dominant |
| base case inpatient pneumonia disutility (-25%) | -356.70 € | 74,694 | 48,483 | Dominant |
| base case inpatient pneumonia disutility (+25%) | -356.70 € | 74,694 | 50,828 | Dominant |
| base case outpatient pneumonia disutility (-25%) | -356.70 € | 74,694 | 49,632 | Dominant |
| base case outpatient pneumonia disutility (+25%) | -356.70 € | 74,694 | 49,678 | Dominant |
| base case IPD herd effects (-25%) | -356.14 € | 74,547 | 49,557 | Dominant |
| base case IPD herd effects (+25%) | -357.29 € | 74,848 | 49,758 | Dominant |
| base case CAP herd effects (-25%) | -356.55 € | 74,671 | 49,642 | Dominant |
| base case CAP herd effects (+25%) | -356.85 € | 74,716 | 49,668 | Dominant |

CAP: all-cause community-aquired pneumania; IPD: invasive pneumococcal disease; LY: life year; QALY: quality adjusted life year
